# Engaging Men Through HIV Self-Testing with Differentiated Care to Improve ART Initiation and Viral Suppression among Men in Malawi (ENGAGE): a study protocol for a randomized control trial

**DOI:** 10.1101/2022.12.22.22283837

**Authors:** Augustine Talumba Choko, Thomas Coates, Misheck Mphande, Kelvin Balakasi, Isabella Robson, Khumbo Phiri, Sam Phiri, Michal Kulich, Michael Sweat, Morna Cornell, Risa Hoffman, Kathryn Dovel

## Abstract

**Background:** Men experience twice the mortality of women while on ART in sub-Saharan Africa (SSA) largely due to late HIV diagnosis and poor retention. Here we propose to conduct an individually randomized control trial (RCT) to investigate the impact of three-month home-based ART (hbART) on viral suppression among men who were not engaged in care.

**Methods and Design:** A programmatic, individually randomized non-blinded, non-inferiority-controlled trial design (ClinicalTrials.org NCT04858243). Through medical chart reviews we will identify “non-engaged” men living with HIV, ≥15years of age who are not currently engaged in ART care, including (1) men who have tested HIV-positive and have not initiated ART within 7 days; (2) men who have initiated ART but are at risk of immediate default; and (3) men who have defaulted from ART. With 1:1 computer block randomization to either hbART or facility-based ART (fbART) arms, we will recruit men from 10-15 high-burden health facilities in central and southern Malawi. The hbART intervention will consist of 3 home-visits in a 3-month period by a certified male study nurse ART provider. In the fbART arm, male participants will be offered counselling at male participant’s home, or a nearby location that is preferred by participants, followed with an escort to the local health facility and facility navigation. The primary outcome is the proportion of men who are virally suppressed at 6-months after ART initiation. Assuming primary outcome achievement of 24.0% and 33.6% in the two arms, 350 men per arm will provide 80% power to detect the stated difference.

**Discussion:** Identifying effective ART strategies that are convenient and accessible for men in SSA is a priority in the HIV world. Men may not (re-)engage in facility-based care due to a myriad of barriers. Two previous trials investigated the impact of hbART on viral suppression in the general population whereas this trial focuses on men. Additionally, this trial involves a longer duration of hbART i.e., three months compared to two weeks allowing men more time to overcome the initial psychological denial of taking ART.

## Introduction

Men are a priority group for HIV services in sub-Saharan Africa (SSA) because of their low uptake of both testing and ART services. In SSA, compared with women, men living with HIV are disproportionately unaware of their HIV status and are not engaged in antiretroviral therapy (ART) programs [1, 2]. Once on ART, men have twice the mortality than women [3, 4], largely due to late diagnosis and poor retention in care [5-8]. Men also cycle through ART programs at a greater rate than women, with high rates of stopping and restarting HIV treatment and multiple extended periods outside HIV care throughout their lifetimes [9, 10]. Once engaged or re-engaged in care, the highest risk for default is within the six several months after (re)initiation [11]. This is a critical period for men to develop the internal motivation and external support needed to sustain ongoing engagement in care, and to learn problem-solving techniques to overcome the multiple barriers facing men. We need to engage more men to achieve the UNAIDS 95/95/95 targets: 95% of people living with HIV are diagnosed, 95% of those diagnosed are on treatment, and 95% of those on treatment are virally suppressed by 2030 [12]. However, there is limited literature on differentiated models of care (DMOC) to support men’s ART initiation (or re-initiation) and early retention in care, and whether such models work for men who have otherwise struggled to engage in care [13-15].

Two overarching barriers prevent HIV-positive men from accessing ART services. First, health facilities usually lack male-friendly services [16-18]. Male-friendly services are private and convenient (requiring minimal time), and offered by health workers who understand the unique needs of men [19-21]. In general, men have little exposure to the health system, which offers little to meet their needs [22, 23], and are unfamiliar with navigating preventative and chronic care services except for outpatient departments [24]. Without male-friendly services men may avoid care. Second, harmful gender norms embedded within health systems and men’s communities de-prioritize men’s health and men’s use of health care [1, 25, 26]. Gender norms that valorize men as strong and self-reliant perpetuate stigma and the fear of unwanted disclosure and can lead men to prioritize short-term benefits such as daily financial earnings and respect from male friends over ART engagement [25, 27, 28].

HIV self-testing (HIVST) users may face additional barriers to engaging in HIV care. HIVST is an effective strategy to reach populations regarded as ‘hard-to-reach’ including men and youth [29, 30]. Recommended by the World Health Organization (WHO), HIVST is being implemented as routine care throughout SSA. However, initial engagement in ART programs may be particularly difficult for HIVST users who have not had to overcome facility-level barriers to testing but now face these barriers when required to initiate ART [31, 32]. The benefits of HIVST (privacy, convenience, and full autonomy) are largely removed when men enter routine ART programs at health facilities. ART engagement must be improved if HIVST is to become a viable option for high-risk groups in SSA [33, 34]. Combining HIV testing strategies with an intervention to increase ART initiation and early retention may support men successfully engaging in HIV care.

Home-based ART (hbART) + male-centered counseling may improve ART (re-)initiation and retention among both HIVST users and traditional testers. Home-based ART for stable clients significantly reduces client time and the cost of accessing services, major deterrents for men still grappling with the decision to (re-)engage in HIV care services [35]. Home-based services also reduce client concerns about privacy and stigma [36, 37] and could be used to facilitate client-centered counseling tailored to the individual man’s needs, which is generally not feasible in busy clinic settings. However, ongoing hbART is expensive for the health system and unsustainable in low-resource settings where the health system is already strained. One alternative is to offer home-based ART (re-)initiation with quick linkage to facility-based ART services. The convenience, privacy, and client-centered nature of three -months of hbART may give men time to accept their status and build coping mechanisms and strategies to overcome barriers to facility-based care.

We propose to conduct an individually randomized control trial in Malawi to test the impact of three-month hbART (plus male-centered counseling and assisted linkage to facility-based ART at the end of this period) on ART (re-)initiation and six-month viral suppression among men who were not engaged in care. The intervention will be compared to standard of care, which includes: a) ART counseling; b) assisted linkage to facility ART; and c) facility-based ART (fbART). We hypothesize that hbART will largely remove barriers to ART initiation and provide coping strategies so that these men can be retained on ART at facilities.

## Materials and Methods

### Objectives

Our primary objective is to test the effect of 3-months of hbART services + male-specific counseling on 6-month viral suppression as compared to standard of care fbART (counseling + assisted facility linkage). Secondary objectives are to: identify individual-, community-, and facility-level factors associated with viral suppression; and to determine the cost and scalability of hbART versus fbART.

### Trial Design

We will use a programmatic, individually randomized non-blinded, non-inferiority-controlled trial design. Men will be randomized 1:1 to either hbART or fbART arms. We will recruit men from 10-15 high-burden health facilities in Malawi.

We developed the study using the SPIRIT (Standard Protocol Items: Recommendations for Interventional Trials) Checklist (see Figure 1 and Supporting Information S1) and the protocol adheres to the SPIRIT recommendations [38]. The protocol (see Supporting Information S2) is approved by the institutional review boards of the University of California, Los Angeles (UCLA) (see Supporting Information S3) and the National Health Sciences Research Council (NHSRC) in Malawi (see Supporting Information S4). The trial is registered with ClinicalTrials.gov as NCT04858243.

**Fig 1.**
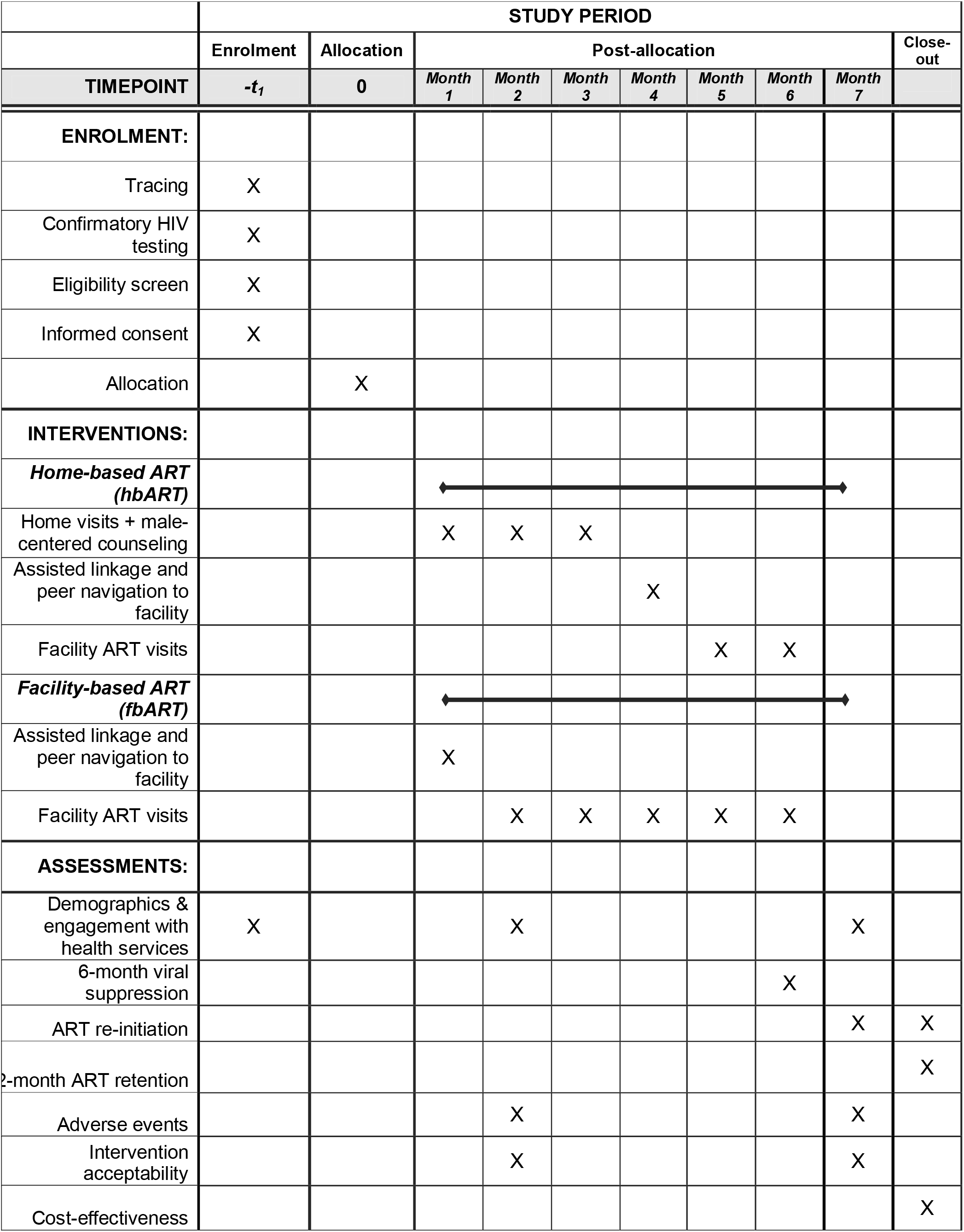
Schedule of enrolment, interventions, and assessments.

### Randomization

Individual men will be block randomized 1:1 to either hbART or fbART study arms using a computer-generated program, with a block size of 3. We will create a randomization list for each participating facility and study ID’s will be linked to the randomization list. Study staff enrolling men will only have access to partial study ID’s (so they are blinded to the randomization order) and will only have access to partial study ID’s for the facility they are actively enrolling at. After enrolment and baseline survey is completed, participants will be assigned a study ID based on the randomization list. Study ID’s will be linked with the pre-assigned blocked randomization, which will already be pre-loaded into the tablet device, but will be unknown to the study staff until survey and randomization modules are completed and saved, ensuring randomization cannot be manipulated or changed by study. Randomization results will appear on the tablet device as a picture on a pre-programmed tablet, and will be shown to the participant in order to maximize transparency and study buy-in.

### Intervention

#### hbART + male-centered counseling

The hbART intervention will consist of 3 home-visits in a 3-month period by a certified <u>male</u> study nurse ART provider. HbART initiation will be scheduled at times convenient for men, including evening and weekends. Men may also choose to initiate in another private location in the community (besides their home). The first home visit will be conducted by a trained nurse and will include confirmatory HIV testing using the Ministry of Health standard algorithm (Determine + Unigold), pre-ART counseling and motivational interviewing, a basic health evaluation, and ART initiation with a 30-day supply of Malawi first-line ART– dolutegravir, tenofovir, and lamivudine as a single tablet (Fig 2). Clients will also be given a 30-day supply of cotrimoxazole, which is standard of care for all HIV-positive individuals.

**Fig 2:**
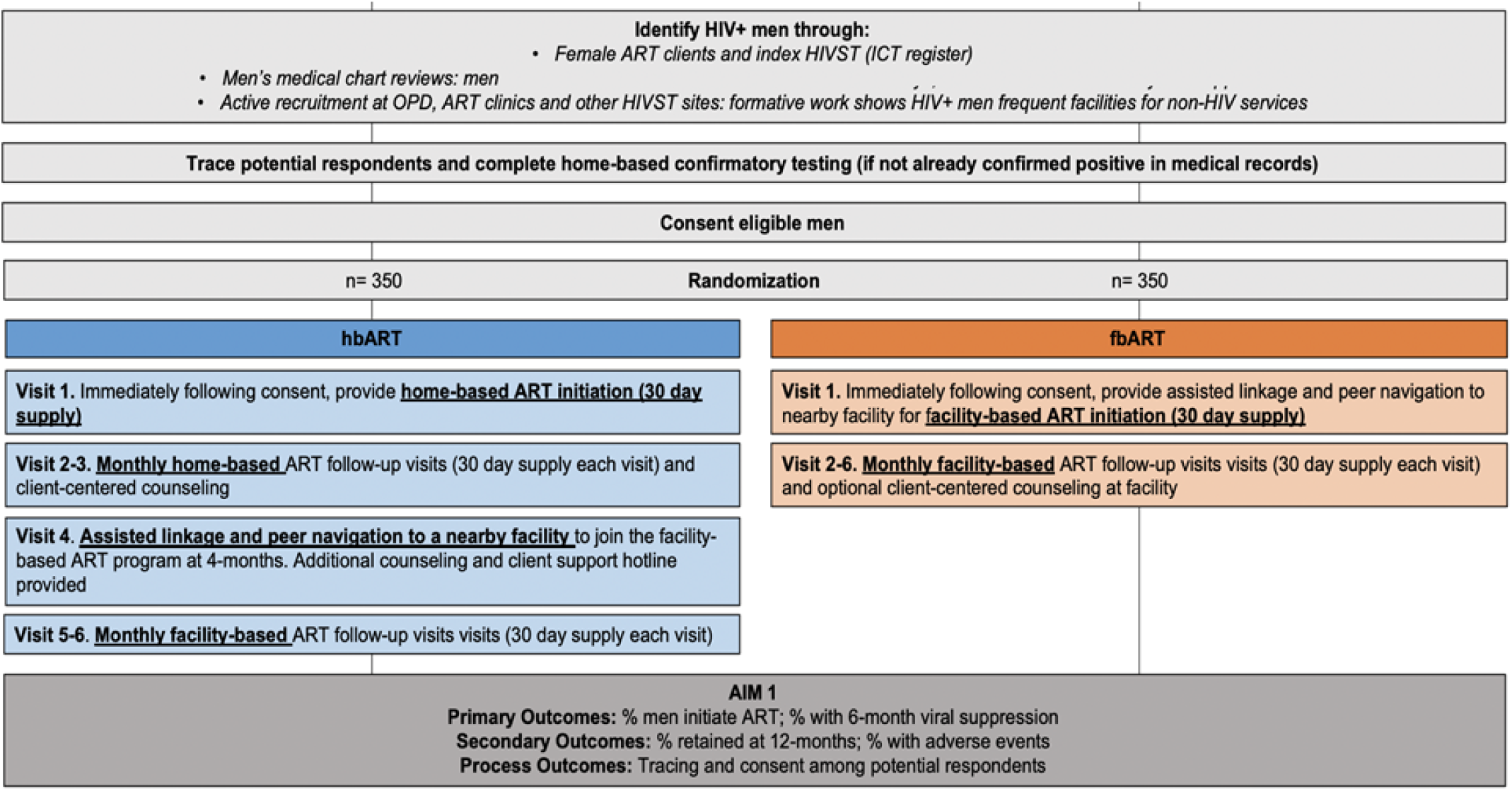
Trial schema.

Prior to ART initiation, the nurse will perform a basic health evaluation to assess WHO staging (see Supporting Information S5)[39]. Any individual WHO stage 3 or 4, or identified as possibly having an active opportunistic infection or other health problem(s) that could complicate hbART, will be referred and escorted to the facility. If the individual stabilizes after facility evaluation, additional ART visits may be performed as hbART. Individuals who refuse home-based services will be given client-centered counseling and the option of being escorted to a local facility for facility-based HIV services. In line with national guidelines, men who refuse ART initiation will receive two additional follow-up visits at one-month intervals in order to assess if they have started ART, and if not, if they would like to do so.

At each hbART visit, motivational interviewing will be performed in preparation for men to engage in facility-based ART services. This includes counseling on the benefits of early ART, strategies for disclosure and positive living, and strategies to overcome facility-based barriers to ART services. Counseling will be adaptive to the needs and concerns of male clients.

At the four-month appointment, the nurse will escort the man to the local health facility to join the facility-based ART cohort. As per national guidelines, individuals will receive monthly facility-based ART appointments that can include counseling by facility staff as needed. Men who wish to attend a facility that is not nearby will be linked with a male counselor from the selected facility. After completing all facility-based ART services for that day, the study nurse will provide additional client-centered counseling to discuss the experience, benefits and challenges associated with facility-based ART, and strategies to overcome barriers.

### Standard of care: fbART

Male participants will be offered counselling at male participant’s home, or a nearby location that is preferred by participants, followed with an escort to the local health facility and facility navigation. The initial home visit will be scheduled at times convenient for men, including evening and weekend hours. Male participants who refuse ART (re-)initiation will receive 3 additional visits in 14-day intervals to offer counselling and encourage initiation.

### Trial setting

The study will be conducted in central and southern Malawi. Malawi’s adult HIV prevalence rate is 8%; there are approximately 330,000 men living with HIV in the country, of which nearly 54,500 are not currently in care and on ART [40]. Men in Malawi live in primarily rural settings, are self-employed, subsistence farmers, the minority have regular access to a private phone, and most are highly mobile.

### Population

Participants will be recruited from between 10 and 15 health facilities in central and southern Malawi that have a high load of HIV+ clients. Different facility types (hospital/health center), management (public/mission), location (rural/urban), and region (central/southern Malawi) will be represented among the included facilities. We will employ medical chart reviews to identify “non-engaged” men living with HIV who are not currently engaged in ART care, including (1) men who have tested HIV-positive and have <u>not initiated</u> ART within 7 days; (2) men who have initiated ART but are at risk of <u>immediate default</u>; and (3) men who have <u>defaulted from ART</u>. Eligibility criteria for men include: (1) ≥15years of age; (2) lives in facility catchment area; and (3) tested HIV-positive and either (a) self-report having not yet initiated ART within 7 days of testing HIV-positive, (b) initiated ART but are at risk of immediate default (i.e., ≥7 days late for their first ART refill appointment after initiation), or (c) initiated ART but have defaulted (i.e., ≥28 days late for any ART refill appointment). Men who have never initiated ART and have no proof of a confirmatory HIV test will be offered a HIV self-test kit prior to enrollment in order to confirm HIV status.

### Study outcomes

The primary study outcome is the proportion of men who are virally suppressed at 6-months after ART initiation. Secondary outcomes include: (1) the proportion of men who (re-)initiate ART; (2) ART retention at 12-months after ART initiation; (3) adverse events experienced (i.e., unwanted disclosure of HIV status, end of relationship, or intimate partner violence); and (4) acceptability of hbART. Process outcomes to be examined include: (1) the proportion of men who were successfully traced; (2) the proportion of eligible men who consented to study participation; (3) men’s exposure to the intervention; and (4) quality of the hbART intervention provided. Viral suppression and retention outcomes will be measured through medical chart reviews. Acceptability of the intervention, adverse events, and exposure and quality of intervention received will be measured through self-reports by male participants during follow-up surveys. All other process outcomes will be measured through study monitoring and evaluation tools.

### Sample size considerations

The study’s sample size was powered to detect differences in viral suppression between the hbART arm and the SOC arm at 6-months. Assuming that 50% of men in the Standard of Care arm and 70% of men in the hbART intervention arm initiate ART, that 60% of men in both arms who initiate ART will achieve viral suppression, and that 20% of men in both arms will be lost to follow-up (and treated as failures for outcome evaluation), the success proportions will be 0.240 and 0.336 in the two arms. The sample size needed to detect this difference with the power of 0.8 is 350 men per arm. The calculation is based on asymptotic normality of log odds ratio. We thus need to enroll and randomize 700 HIV-positive men.

### Data Collection

#### Recruitment

Men who have tested positive for HIV but are not engaged in HIV services will be identified as potential study participants through medical chart reviews as well as in-person recruitment at study sites. The medical chart reviews will involve reviewing three different types of medical charts and registers to identify (1) men who have tested positive for HIV and have not initiated ART; (2) men who have initiated ART but subsequently missed their 4-week appointment (≥7 days late); and (3) men who have defaulted from ART (≥28 days late for any other appointment after the first 4-week follow-up). We will also conduct in-person screening and recruitment of men at outpatient departments (OPDs) since previous research indicates that Malawian men frequently seek care at outpatient departments (OPDs), including men who have defaulted from ART but seek curative care at OPDs.[24] Finally, we will screen and recruit women living with HIV who attend ART clinics to asses if they have male partner who is potentially eligible for the study. Women with potentially eligible partners will be given a Study Invitation Card to give to her male partner.

#### Tracing and Eligibility Screening

The study team will trace individuals identified through medical chart reviews. Three attempts will be made to trace each potential participant before they are considered lost to follow-up. All screening and enrollment will take place in-person, although phones will be used for an initial screen if a potential participant’s phone number is available. Immediately after being traced, men will complete an eligibility screen in a quiet, private, and convenient location. Female ART clients identified as having potentially eligible male partners will also be traced and asked to complete a screen for her partner; if the male partner may be eligible, the female ART client will be told about the study and asked to have her male partner attend an in-person meeting or to share a Study Invitation Card with him.

#### Consent, Enrollment, and Baseline Survey

Written informed consent will be obtained from men who are found to be eligible for the study during screening and interested in participating. Following Malawian ethical protocols, written informed consent will be obtained from parents and/or guardians of adolescents <18 years of age. Assent will also be required to demonstrate their individual understanding of study participantion. After the informed consent procedure is complete, participants will be immediately enrolled in the study and will then complete a baseline survey that gathers key demographic data (i.e., marital status, number of children, employment, self-rated health) and previous engagement with HIV and non-HIV health services. Surveys will be conducted face-to-face in the local language (Chichewa) by trained research assistants; surveys will be programmed and responses recorded on electronic tablets using SurveyCTO software (http://www.surveycto.com). Following completion of the baseline survey, men will be block randomized 1:1 to either the home-based ART arm or the SOC arm. Based on their assigned arm, men will be subsequently referred to the appropriate HCW for intervention implementation.

#### Intervention Implementation

Health care workers will implement all intervention activities. Standard of care fbART will be implemented by routine facility staff. Trained study nurses will implement all activities in the hbART arm. All HCWs will be based at participating health facilities and will also have routine duties within the facility, increasing the replicability of the interventions in real-world settings.

#### Follow-Up Data

Research Assistants will administer follow-up surveys at 2- and 7-months after enrollment. Follow-up surveys will measure exposure to (and acceptability of) the interventions, changes in key demographics since enrollment (i.e., marital status, number of children, employment, self-rated health), any adverse events since enrollment (i.e., unwanted status disclosure, termination of relationship due to the intervention), and experiences with ART services. Follow-up surveys will be conducted at participant’s home or at any location and time that is convenient for the participant.

Medical chart reviews will be conducted to assess men’s engagement with ART services. Individuals who cannot be found through medical chart review will be followed-up in person to confirm retention outcome using their personal medical records (i.e., did men transfer to another facility, was their poor documentation at the health facility, or did men disengage in care). Men who cannot be reached or are lost to follow-up in any arm will be counted as failures for that specific ART outcome of interest.

### Cost data

We will conduct an incremental cost-effectiveness analysis and mathematical modelling to determine national scale-up potential. The average cost per successful outcome (early ART retention) will be calculated and compared across arms incrementally.

Costs will be measured from the health care provider. We will use micro-costing methods by first creating an inventory of all the resources used to achieve the observed study outcomes including:

- Male-focused counseling interactions (staff cadre, training received, duration of interaction and distance from facility travelled where applicable)
- Peer support interactions (staff cadre, training received, duration of interaction and distance from facility travelled where applicable)
- Provider interactions (staff cadre, training received, duration of interaction and distance from facility travelled where applicable)

For each study patient, the quantity (number of units) of resources used will be determined. Unit costs of resources, which are not human subject data, will be obtained from external suppliers and the site’s finance and procurement records and multiplied by the resource usage data to provide an average cost per study patient across centers in each study arm.

### Analysis plan

Data analysis will be conducted in Stata 14 (Stata Corp., TX, USA). We will use the Consolidated Standards of Reporting Trials (CONSORT) standards for reporting trial outcomes [41]. Using an intention-to-treat analysis, all randomized men will be included in the analysis of primary outcomes; men with missing outcome assessment due to loss to follow-up will be treated as outcome failures. All primary outcomes are binary. We will calculate descriptive statistics, including mean/median, variation (standard deviation, kurtosis), range, and frequency distributions for the demographic and clinical characteristics, overall and by study arm. Logistic regression models will be developed for the probability of a positive outcome, with sociodemographic factors included as covariates in a suitable form (linear/spline/factor). Differences in the prevalence of each of the outcomes of interest will be examined by study arm as well as by other factors of interest including demographic characteristics (e.g., age), couple characteristics, and knowledge/perceptions and biomedical knowledge. The differences will be evaluated using t-tests, Mann-Whitney U test (or other non-parametric tests), chi-square methods, and Fisher’s exact test as appropriate.

We recognize that men’s consent to participate in the study may bias the sample enrolled in the study. As such, we will also conduct sensitivity analyses to account for men who we were never able to contact (unreachable men) and men who refused to participate in the full trial (refusers). Data on unreachable men will be collected via their female partner using a brief survey on male and female demographics, sexual partnerships and couple dynamics, and men’s history with health services, and HIV services specifically (as reported by female partners). Refusers will be offered a one-time survey immediately following refusal for the larger study. The same data will be collected, as described above.

### Cost-Effectiveness

Using the average cost per patient as described above, we will then estimate the cost per outcome achieved in each arm. The main measure of effectiveness for the cost-effectiveness analysis will be both the primary study outcome (viral suppression at 6-months). We will calculate the difference in cost divided by the difference in effectiveness among study arms. Costs will be reported as means (standard deviations) and medians (IQRs) in USD, using the exchange rate prevailing during the follow up period.

### National scale-up modeling

To determine the budget impact and affordability of the intervention arms, we will parameterize a national scale-up model using the study output. To determine the total cost and impact of the three intervention arms, as well as combinations of interventions, we will model cost and impact out to early ART retention (ART initiation <u>and</u> completion of the 4-week ART refill appointment). The following parameters to be estimated from this trial include:

- Proportion of men that initiate ART
- Proportion of men that complete the 4-week ART refill appointment
- Proportion of men that reached viral suppression at 6-months after ART initiation
- Proportion of men that are retained in care at 12-months after ART initiation

We will then estimate the expected increase in the number of men linked to ART after index HIVST, adjusted by facility type where possible, by each intervention arm. The number of facility-level HIV tests conducted through index testing at all 652 public healthcare facilities in Malawi from Oct 2019-Sept 2020 will be used for these national calculations. Each intervention will be tested separately in this model, as well as different combination of interventions.

Different scenarios will be explored where interventions are used at different facilities (urban versus rural targeting of interventions, geospatial targeting of interventions), or different groups of men within the same facility (where data suggest that different demographics of men respond differently to the different interventions).

The national-level costs and expected number of men linked to ART, by each intervention and combinations of interventions, will be reported from this model. We will then contextualize the national cost of each intervention with a short-term 3-year budget impact: percent increase (or decrease) of the national HIV treatment budget with the inclusion of one of these interventions.

## Discussion

Identifying effective ART strategies that are convenient and accessible for men in SSA is a priority in the HIV world. Men may not (re-)engage in facility-based care due to (1) fear of unwanted disclosure and stigma due to <u>lack of privacy</u>; (2) <u>time/cost required</u> to access care; (3) <u>poor knowledge</u> about the benefits of early ART initiation or how to navigate the health system [42]. The immediate costs associated with engaging in an ART program must be decreased, and men counseled around the fact that immediate costs are outweighed by immediate gains (treatment as prevention), as well as long-term gains (better health outcomes and potential for income generation).

In the Engage trial, we aim to investigate the effect of home-based ART (hbART) on men’s ART initiation and viral suppression. Home-based ART is a differentiated service delivery (DSD) model and a variant of community-based ART. Notable previous randomized trials have showed that home-based ART initiation with two weeks starter pack of ART at home increased demand for ART in Malawi [43] and in Lesotho [44]. Both of these two previous trials involved the general population and not men specifically. as is the case in the present trial. Given the myriad of barriers that men face – mostly owing to concerns around access and privacy of services [42] – we posit that the hbART strategy will be effective among men. Our trial also involves a longer duration of home-based ART of three months compared to two weeks. We theorize that this component may allow men more time to overcome the initial psychological denial of taking ART, and develop strategies to overcome internal and external barriers to care, before accessing the health facility for continuation.

Other than investigating the effect of the hbART, we are keen to assess the costs and cost-effectiveness of this intervention. Additionally, more attention will be given to the presence of any adverse outcomes including unwanted status disclosure. Understanding these adverse outcomes is key to identifying aspects of the intervention that may require further refinement before being taken to scale.

There are some potential limitations in this study. Firstly, the nature of home-based ART dispensation requires that clients are clinically stable. This implies that although the hbART may be found to be effective at increasing ART initiation and viral suppression it may not apply to all HIV positive men. Secondly, there is a chance that the implementation of hbART via a study nurse may be associated with stigma or unwanted disclosure in the community [43]. Finally, the intervention implementation may become logistically difficult to implement for a mobile population where home visits imply physical interaction with the implementation team.

Men continue to be underrepresented in ART programs throughout SSA. Through the Engage trial, we will engage men living with HIV but not currently in care, and test the impact of home-based care for three months on men’s 6-month retention in ART programs and 6-month viral suppression. Such research focused specifically on men promises to help improve the gender imbalances historically present in HIV programs in the region.

## Data Availability

This protocol manuscript uses openly available data found in the reference section. All data for the trail will be made available following its completion.

## Authors’ contribution

All authors contributed to study design and writing of the paper. Conceived of the study: TC and KD. Provided statistical expertise in trial design: MK and MS. Provided expertise in interventional design: AC, TC, KB, KP, MC, RH, and KD. Implemented the trial: MM and IR. Conducting primary statistical analysis: KB and MK. Conducting primary cost-effectiveness analysis: MS. Responsible for first draft: TC and KD. All authors read and approved the final manuscript.

## Acknowledgements

We wish to thank the Malawi Ministry of Health for their support of this trial. We would also like to acknowledge Agnes Moses, Elijah Chikuse, and Joep J can Oosterhout for their contributions to protocol development and for their support of study implementation.

## Supporting information

File S1. SPIRIT Checklist.

File S2. IRB-approved study protocol

File S3. IRB-approval letter (UCLA)

File S4. IRB-approval letter (Malawi)

File S5. WHO clinical staging of HIV disease in adults, adolescents and children

File S6. Funding Letter

