## Supporting Information S4 for "Engaging Men Through HIV Self-Testing with Differentiated Care to Improve ART Initiation and Viral Suppression among Men in Malawi (ENGAGE): a study protocol for a randomized control trial"

Telephone: + 265 789 400
Facsimile: + 265 789 431

**All Communications should be addressed to: The Secretary for Health**

*In reply please quote No.* .......................
MINISTRY OF HEALTH
P.O. BOX 30377
LILONGWE 3
MALAWI

**27th May, 2022**

**Dr. Kathryn Dovel**
Partners in Hope

Dear Sir/Madam

**RE: Protocol #20/07/2562: Engaging Men through HIV Self-Testing with Differentiated Care for ART Initiation and Viral Suppression among Men in Malawi**

Thank you for the above titled proposal that you submitted to the National Health Sciences Research Committee (NHSRC) for review. Please be advised that the NHSRC has **reviewed** and **approved the continuation** of the above named study.

- **APPROVAL NUMBER : 2562**
- The above details should be used on all correspondences, consent forms and documents as appropriate.
- **APPROVAL DATE :27/05/2022**
- **EXPIRATION DATE :26/05/2023**
  This approval expires on **26/05/2023**. After this date, this project may only continue upon renewal. For purposes of renewal, a progress report on a standard form obtainable from the NHSRC Secretariat should be submitted one month before the expiration date for continuing review.
- **SERIOUS ADVERSE EVENT REPORTING:** All serious problems having to do with subject safety must be reported to the NHSRC within 10 working days using standard forms obtainable from the NHSRC Secretariat.
- **MODIFICATIONS**: Prior NHSRC approval using forms obtainable from the NHSRC Secretariat is required before implementing any changes in the protocol (including changes in the consent documents). You may not use any other consent documents besides those approved by the NHSRC.
- **TERMINATION OF STUDY**: On termination of a study, a report has to be submitted to the NHSRC using standard forms obtainable from the NHSRC Secretariat.
- **OTHER**: Please be reminded to send in copies of your final research results for our records (Health Research Database).

Kind regards from the NHSRC Secretariat.

National Health Sciences Research Committee
2022 -05- 27
APPROVED

..........................................................................

For: **CHAIRPERSON, NATIONAL HEALTH SCIENCES RESEARCH COMMITTEE**
Promoting Ethical Conduct of Research [1]

**Executive Committee:** *Dr. Martias Joshua (Chairperson), Dr. Evelyn Chitsa Banda (Vice-Chairperson)*
**Registered withthe USA Office for Human Research Protections (OHRP) as an International IRBIRB Number IRB00003905 FWA00005976**
