## Supporting Information S5 for "Engaging Men Through HIV Self-Testing with Differentiated Care to Improve ART Initiation and Viral Suppression among Men in Malawi (ENGAGE): a study protocol for a randomized control trial"

**Supporting Information: S5** **WHO clinical staging of HIV disease in adults, adolescents and children**

Source: Adapted from WHO case definitions of HIV for surveillance and revised clinical staging and immunological classification of HIV-related disease in adults and children. Geneva, World Health Organization, 2007 (www.who.int/hiv/pub/guidelines/ HIVstaging150307.pdf).

| **Adults and adolescents^a^** | **Children** |
| --- | --- |
| **Clinical stage 1** | |
| Asymptomatic  Persistent generalized lymphadenopathy | Asymptomatic  Persistent generalized lymphadenopathy |
| **Clinical stage 2** | |
| Moderate unexplained weight loss (<10% of presumed or measured body weight)  Recurrent respiratory tract infections (sinusitis, tonsillitis, otitis media, pharyngitis)  Herpes zoster  Angular cheilitis  Recurrent oral ulceration  Papular pruritic eruption  Fungal nail infections  Seborrhoeic dermatitis | Unexplained persistent hepatosplenomegaly  Recurrent or chronic upper respiratory tract infections  (otitis media, otorrhoea, sinusitis, tonsillitis)  Herpes zoster  Lineal gingival erythema  Recurrent oral ulceration  Papular pruritic eruption  Fungal nail infections  Extensive wart virus infection  Extensive molluscum contagiosum  Unexplained persistent parotid enlargement |
| **Clinical stage 3** | |
| Unexplained severe weight loss (>10% of presumed or measured body weight)  Unexplained chronic diarrhoea for longer than 1 month  Unexplained persistent fever (intermittent or constant for longer than 1 month)  Persistent oral candidiasis  Oral hairy leukoplakia  Pulmonary tuberculosis  Severe bacterial infections (such as pneumonia, empyema, pyomyositis, bone or joint infection, meningitis, bacteraemia)  Acute necrotizing ulcerative stomatitis, gingivitis or periodontitis  Unexplained anaemia (<8 g/dl), neutropaenia (<0.5 x 10^9^/l) and/or chronic thrombocytopaenia (<50 x 10^9^/l) | Unexplained moderate malnutrition^b^ not adequately responding to standard therapy  Unexplained persistent diarrhoea (14 days or more)  Unexplained persistent fever (above 37.5°C, intermittent or constant, for longer than one 1 month)  Persistent oral candidiasis (after first 6 weeks of life)  Oral hairy leukoplakia  Lymph node tuberculosis  Pulmonary tuberculosis  Severe recurrent bacterial pneumonia  Acute necrotizing ulcerative gingivitis or periodontitis  Unexplained anaemia (<8 g/dl), neutropaenia  (<0.5 x 10^9^/l) or chronic thrombocytopaenia (<50 x 10^9^/l) |
| **Adults and adolescents^a^** | **Children** |
| **Clinical stage 3** | |
|  | Symptomatic lymphoid interstitial pneumonitis  Chronic HIV-associated lung disease, including bronchiectasis |
| **Clinical stage 4^c^** | |
| HIV wasting syndrome  Pneumocystis (jirovecii) pneumonia  Recurrent severe bacterial pneumonia  Chronic herpes simplex infection (orolabial, genital or anorectal of more than 1 month’s duration or visceral at any site)  Oesophageal candidiasis (or candidiasis of trachea, bronchi or lungs) Extrapulmonary tuberculosis  Kaposi sarcoma  Cytomegalovirus infection (retinitis or infection of other organs)  Central nervous system toxoplasmosis  HIV encephalopathy  Extrapulmonary cryptococcosis, including meningitis  Disseminated nontuberculous mycobacterial infection  Progressive multifocal leukoencephalopathy  Chronic cryptosporidiosis  Chronic isosporiasis  Disseminated mycosis (extrapulmonary histoplasmosis, coccidioidomycosis)  Lymphoma (cerebral or B-cell non-Hodgkin)  Symptomatic HIV-associated nephropathy or cardiomyopathy  Recurrent septicaemia (including nontyphoidal Salmonella)  Invasive cervical carcinoma  Atypical disseminated leishmaniasis | Unexplained severe wasting, stunting or severe malnutrition^d^ not responding to standard therapy  Pneumocystis (jirovecii) pneumonia  Recurrent severe bacterial infections (such as empyema, pyomyositis, bone or joint infection, meningitis, but excluding pneumonia)  Chronic herpes simplex infection (orolabial or cutaneous of more than 1 month’s duration or visceral at any site)  Oesophageal candidiasis (or candidiasis of trachea, bronchi or lungs)  Extrapulmonary tuberculosis  Kaposi sarcoma  Cytomegalovirus infection (retinitis or infection of other organs with onset at age more than 1 month)  Central nervous system toxoplasmosis (after the neonatal period) HIV encephalopathy  Extrapulmonary cryptococcosis, including meningitis  Disseminated nontuberculous mycobacterial infection  Progressive multifocal leukoencephalopathy  Chronic cryptosporidiosis (with diarrhoea)  Chronic isosporiasis  Disseminated endemic mycosis (extrapulmonary histoplasmosis, coccidioidomycosis, penicilliosis) Cerebral or B-cell non-Hodgkin lymphoma  HIV-associated nephropathy or cardiomyopathy |

a In the development of this table, adolescents were defined as 15 years or older. For those aged less than 15 years, the clinical

staging for children should be used.

b For children younger than 5 years, moderate malnutrition is defined as weight-for-height <–2 z-score or mid-upper arm

circumference ≥115 mm to <125 mm.

c Some additional specific conditions can be included in regional classifications, such as penicilliosis in Asia, HIV-associated rectovaginal fistula in southern Africa and reactivation of trypanosomiasis in Latin America. ^d^ For children younger than 5 years of age, severe wasting is defined as weight-for-height <–3 z-score; stunting is defined as length-for-age/height-for-age <–2 z-score; and severe acute malnutrition is either weight for height <–3 z-score or mid-upper arm circumference <115 mm or the presence of oedema.
