## Supporting Information S for "Engaging Men Through HIV Self-Testing with Differentiated Care to Improve ART Initiation and Viral Suppression among Men in Malawi (ENGAGE): a study protocol for a randomized control trial"

### University of California, Los Angeles
### Award Snapshot

#### Section I: Award Summary

| | | | |
|---|---|---|---|
| **Principal Investigator:** | Coates, Thomas J | **Fund Number:** | 30467 |
| **Sponsor:** | NIH-NIMH National Institute of Mental Health [000052] | **Sponsor Award Number:** | 5R01MH122308-03 |
| **Administering Unit:** | MEDICINE-INFECTIOUS DISEASE [1560] | **Prime Sponsor:** | N/A |
| **Project Title:** | Engaging Men Through HIV Self-Test and Differentiated Care Models to Increase ART Initiation and Viral Suppression in Malawi | | |
| | | **Current Action:** | Continuation |
| **Current Budget Period:** | 6/1/2022 - 5/31/2023 | **Funds Awarded this Action:** | $515,387 |
| **Project Period:** | 6/17/2020 - 5/31/2025 | **Total Funds Awarded to Date:** | $1,676,685 |

- ***See Section VIII for Other Investigators***
- ***For a History of Actions on this award, refer to the Award Snapshot Attachment***

#### Section II: Special Attention Needed

1. Changes in the status of the Principal Investigator or other key personnel on the award require prior approval from the Sponsor. Requests for prior approval must be processed through OCGA. Notify OCGA in advance or as soon as you become aware of any changes (or anything requiring prior approval).
2. This award is subject to a sponsor salary cap limitation. Salary Cap Type: Health and Human Services
3. Review the Award Snapshot Attachment and the Award document for additional terms and conditions.

#### Section III: Award Demographics

| | | | |
|---|---|---|---|
| **Sponsor Award Number:** | 5R01MH122308-03 | **UCLA PATS Number:** | 20200758 |
| **Proposal Type:** | Resubmission - New | **Award Type:** | Grant |
| **Program Type:** | Applied Org Research | **Special Program Type:** | Not applicable |
| **Award Status:** | Awarded/Fully Executed | **Location:** | On Site |
| **Special Payment Type:** | None | **Pre-Award Spend:** | 90 days, 03/19/2020 |

| Budget Period | Transaction Budget Period | Direct Costs | F&A Costs | Total | F&A Rate | F&A Base | Payment Basis | Award Status | Action Type |
|---|---|---|---|---|---|---|---|---|---|
| 1 | 06/17/2020 - 05/31/2021 | $475,568 | $104,570 | $580,138 | 56.0 % | MTDC | Cost Reimb | Awarded/Fully Executed | New |
| 1 | 06/17/2020 - 05/31/2021 | $52,840 | $10,063 | $62,903 | 56.0 % | MTDC | Cost Reimb | Awarded/Fully Executed | Modification/Amendment |
| 2 | 06/01/2021 - 05/31/2022 | $438,590 | $79,667 | $518,257 | 56.0 % | MTDC | Cost Reimb | Awarded/Fully Executed | Continuation |
| 3 | 06/01/2022 - 05/31/2023 | $438,583 | $76,804 | $515,387 | 56.0 % | MTDC | Cost Reimb | Awarded/Fully Executed | Continuation |
| 4 | 06/01/2023 - 05/31/2024 | $438,583 | $76,804 | $515,387 | 56.0 % | MTDC | Cost Reimb | Anticipated/Committed | Continuation |
| 5 | 06/01/2024 - 05/31/2025 | $461,707 | $82,329 | $544,036 | 56.0 % | MTDC | Cost Reimb | Anticipated/Committed | Continuation |

#### Section IV: Subawards Approved in the Award

| Subawardee | Budget Period |
|---|---|
| Charles University in Prague (Univerzita Karlova V Praze DBA) (Czech Republic)<br>Medical University of South Carolina<br>Partners in Hope (PIH) (Malawi) | 06/17/2020 - 05/31/2021 |
| Charles University in Prague (Univerzita Karlova V Praze DBA) (Czech Republic)<br>Medical University of South Carolina<br>Partners in Hope (PIH) (Malawi) | 06/01/2021 - 05/31/2022 |
| Charles University in Prague (Univerzita Karlova V Praze DBA) (Czech Republic)<br>Medical University of South Carolina<br>Partners in Hope (PIH) (Malawi) | 06/01/2022 - 05/31/2023 |

#### Section V: Training Grant Approved Slots

#### Section VI: Program Income, Cost Sharing and Compliance Requirements

| Anticipated Program Income | Anticipated Program Income Type |
|---|---|
| No | - |

| Mandatory Cost Sharing? | Unfunded Effort (other than salary over the cap) | Amount |
|---|---|---|
| No | No | |

| Special Review Type | Approval Status | Reference |
|---|---|---|
| Terms and Conditions | Approved | YR1: Lift IRB Restriction |
| Human Subjects | See System of Record | 21-000209 |

#### Section VII: Deliverables

As you prepare the required reporting/deliverable to the Sponsor for this project keep in mind that it may contain patentable information. The TDG Technology Transfer Officers are ready to meet or speak with you to discuss your pending work and you are encouraged to report potential inventions at any and all stages of your research. Invention disclosures can be submitted to http://tdg.ucla.edu.submit-invention-report and upon receipt TDG will be in touch with you to discuss your work. **Note that filing a technical report without consulting TDG may jeopardize UCLA's ability to secure a patent to protect your work.**

***Non-Financial Deliverables:***

| Deliverable Category | Frequency | Type | Due Date | Status |
|---|---|---|---|---|
| Tech/Scientific | Annual | Progress Report | 04/15/2021 | Submitted |
| Tech/Scientific | Annual | Progress Report | 04/15/2022 | Past Due |
| Tech/Scientific | Annual | Progress Report | 04/15/2023 | Not Started |
| Tech/Scientific | Annual | Progress Report | 04/15/2024 | Not Started |
| Invention/Patent | One Time | Final | 09/28/2025 | Not Started |
| Tech/Scientific | One Time | Final | 09/28/2025 | Not Started |

***Financial Deliverables:***

| Deliverable Category | Frequency | Type | Due Date | Status |
|---|---|---|---|---|
| Financial Report | Once | Final | 09/28/2025 | Not Started |

#### Section VIII: Other Investigators

| Role | Name |
|---|---|
| PD/PI | Coates, Thomas J |

#### Section IX: Contacts

| Contacts | |
|---|---|
| OCGA | Pak, Jessica |
| EFM | Hwang, Ruth Sue |

**UCLA PATS NUMBER:** 20200758

**Alert(s)**

1. Please review and adhere to the award terms and conditions.
2. Research and Development (R&D): All awards issued by the National Institutes of Health (NIH) meet the definition of "Research and Development" at 45 CFR Part§ 75.2**.** As such, auditees should identify NIH awards as part of the R&D cluster on the Schedule of Expenditures of Federal Awards (SEFA).
3. **[LIFTED]** RESTRICTION - HUMAN SUBJECTS: This award has been issued without a currently valid certification of IRB approval for one or more protocol(s). Only activities that are clearly severable and independent from activities that involve human subjects or activities for which IRB approval has been obtained may be conducted. Activities for the protocol(s) cited above may not be conducted until IRB approval of the protocol(s).
4. **PARTICIPANT RECRUITMENT – MILESTONES**: Future NIMH support for this study is contingent upon adequate participant recruitment based on projected milestones as approved in the Recruitment Milestone Reporting system (RMR) on 6/15/2021. It is expected that **580** of the **1100** total projected participants will be recruited by 4/1/2023.

**Reference Document(s)**

1. Award(s) available via the ORA Award Status & Snapshot Report [http://portal.research.ucla.edu/index.aspx?Section=PostAward]
2. NIH Grants Policy Statement December 2021[ https://grants.nih.gov/policy/nihgps/index.htm] Effective for all NIH grants and cooperative agreements with budget periods beginning on or after December 1, 2021.
3. Federal-Wide Research Terms and Conditions November 2020 https://www.nsf.gov/awards/managing/rtc.jsp
    a. RTC Prior Approval and Other Requirements Matrix November 2020
    b. RTC NIH Agency-Specific Requirements November 2020
4. 2 CFR 200: Uniform Administrative Requirements for Awards and Subawards to Institutions of Higher Education, Hospitals, Other Nonprofit Organizations, and Commercial Organizations, Revised November 2020
5. **AWARD NOTICE:** This award has been made in response to the application submitted under the Funding Opportunity Announcement PA19-042 which can be referenced at: https://grants.nih.gov/grants/guide/pa-files/PA-19-042.html

**Action(s)**

1. Sponsor award dated 06/17/2020 provides funding in the amount of $580,138 for Year 1.
2. Sponsor award dated 09/15/2020 provides additional funding in the amount of $62,903 for Year 1. This award is revised to restore administrative funding cuts to year 01.
3. Sponsor award dated 06/02/2021 revises the award to reflect an acceptance of the certification of IRB approval.
4. Sponsor award dated 06/02/2021 provides continuation funding in the amount of $518,257 for Year 2.
5. *This Snapshot*: Sponsor award dated 05/10/2022 provides continuation funding in the amount of $515,387 for Year 3.

**Department of Health and Human Services**
National Institutes of Health
NATIONAL INSTITUTE OF MENTAL HEALTH

**Notice of Award**
FAIN# R01MH122308
**Federal Award Date**
05/10/2022

#### Recipient Information

**1. Recipient Name**
UNIVERSITY OF CALIFORNIA, LOS ANGELES
10889 WILSHIRE BLVD STE 700

LOS ANGELES, 90024

**2. Congressional District of Recipient**
33

**3. Payment System Identifier (ID)**
1956006143A1

**4. Employer Identification Number (EIN)**
956006143

**5. Data Universal Numbering System (DUNS)**
092530369

**6. Recipient's Unique Entity Identifier**
RN64EPNH8JC6

**7. Project Director or Principal Investigator**
Thomas J. Coates, PHD
Professor

310-557-3044

**8. Authorized Official**
FRANK FALCONII

310-206-9898

#### Federal Agency Information

**9. Awarding Agency Contact Information**
Jackie Chia
Grants Management Representative
NATIONAL INSTITUTE OF MENTAL HEALTH

301-443-1341

**10. Program Official Contact Information**
Gregory Greenwood
Program Officer
NATIONAL INSTITUTE OF MENTAL HEALTH

(240) 669-5532

#### Federal Award Information

**11. Award Number**
5R01MH122308-03

**12. Unique Federal Award Identification Number (FAIN)**
R01MH122308

**13. Statutory Authority**
42 USC 241 42 CFR 52

**14. Federal Award Project Title**
Engaging men through HIV self-test and differentiated care models to increase ART initiation and viral suppression in Malawi

**15. Assistance Listing Number**
93.242

**16. Assistance Listing Program Title**
Mental Health Research Grants

**17. Award Action Type**
Non-Competing Continuation

**18. Is the Award R&D?**
Yes

##### Summary Federal Award Financial Information

| | |
|---|---|
| **19. Budget Period Start Date** 06/01/2022 **– End Date** 05/31/2023 | |
| **20. Total Amount of Federal Funds Obligated by this Action** | $515,387 |
| 20 a. Direct Cost Amount | $438,583 |
| 20 b. Indirect Cost Amount | $76,804 |
| **21.** Authorized Carryover | |
| **22.** Offset | |
| **23.** Total Amount of Federal Funds Obligated this budget period | $515,387 |
| **24. Total Approved Cost Sharing or Matching, where applicable** | $0 |
| **25. Total Federal and Non-Federal Approved this Budget Period** | $515,387 |
| ---------- |  |
| **26. Project Period Start Date** 06/17/2020 **– End Date** 05/31/2025 | |
| **27.** Total Amount of the Federal Award including Approved Cost Sharing or Matching this Project Period | $1,676,685 |

**28. Authorized Treatment of Program Income**
Additional Costs

**29. Grants Management Officer - Signature**
Jackie Chia

**30. Remarks**
Acceptance of this award, including the "Terms and Conditions," is acknowledged by the recipient when funds are drawn down or otherwise requested from the grant payment system.

*RESEARCH*
Department of Health and Human Services
National Institutes of Health

NATIONAL INSTITUTE OF MENTAL HEALTH

**SECTION I – AWARD DATA – 5R01MH122308-03**

**Principal Investigator(s):**
Thomas J. Coates, PHD

Dear Authorized Official:

The National Institutes of Health hereby awards a grant in the amount of $515,387 (see “Award Calculation” in Section I and “Terms and Conditions” in Section III) to UNIVERSITY OF CALIFORNIA LOS ANGELES in support of the above referenced project. This award is pursuant to the authority of 42 USC 241 42 CFR 52 and is subject to the requirements of this statute and regulation and of other referenced, incorporated or attached terms and conditions.

Acceptance of this award, including the "Terms and Conditions," is acknowledged by the recipient when funds are drawn down or otherwise requested from the grant payment system.

Each publication, press release, or other document about research supported by an NIH award must include an acknowledgment of NIH award support and a disclaimer such as “Research reported in this publication was supported by the National Institute Of Mental Health of the National Institutes of Health under Award Number R01MH122308. The content is solely the responsibility of the authors and does not necessarily represent the official views of the National Institutes of Health.” Prior to issuing a press release concerning the outcome of this research, please notify the NIH awarding IC in advance to allow for coordination.

Award recipients must promote objectivity in research by establishing standards that provide a reasonable expectation that the design, conduct and reporting of research funded under NIH awards will be free from bias resulting from an Investigator’s Financial Conflict of Interest (FCOI), in accordance with the 2011 revised regulation at 42 CFR Part 50 Subpart F. The Institution shall submit all FCOI reports to the NIH through the eRA Commons FCOI Module. The regulation does not apply to Phase I Small Business Innovative Research (SBIR) and Small Business Technology Transfer (STTR) awards. Consult the NIH website http://grants.nih.gov/grants/policy/coi/ for a link to the regulation and additional important information.

If you have any questions about this award, please direct questions to the Federal Agency contacts.

Sincerely yours,

Jackie Chia
Grants Management Officer
NATIONAL INSTITUTE OF MENTAL HEALTH

Additional information follows

Cumulative Award Calculations for this Budget Period (U.S. Dollars)

| | |
|---|---|
| **Salaries and Wages** | $91,521 |
| **Fringe Benefits** | $25,255 |
| **Personnel Costs (Subtotal)** | $116,776 |
| **Materials & Supplies** | $1,800 |
| **Travel** | $9,270 |
| **Other** | $325 |
| **Subawards/Consortium/Contractual Costs** | $306,812 |
| **Publication Costs** | $3,600 |

| | |
|---|---|
| **Federal Direct Costs** | $438,583 |
| **Federal F&A Costs** | $76,804 |
| **Approved Budget** | $515,387 |
| **Total Amount of Federal Funds Authorized (Federal Share)** | $515,387 |
| **TOTAL FEDERAL AWARD AMOUNT** | $515,387 |
| **AMOUNT OF THIS ACTION (FEDERAL SHARE)** | $515,387 |

| SUMMARY TOTALS FOR ALL YEARS (for this Document Number) | | |
|---|---|---|
| YR | THIS AWARD | CUMULATIVE TOTALS |
| 3 | $515,387 | $515,387 |
| 4 | $515,387 | $515,387 |
| 5 | $544,036 | $544,036 |

Recommended future year total cost support, subject to the availability of funds and satisfactory progress of the project

**Fiscal Information:**

| | |
|---|---|
| **Payment System Identifier:** | 1956006143A1 |
| **Document Number:** | RMH122308A |
| **PMS Account Type:** | P (Subaccount) |
| **Fiscal Year:** | 2022 |

| IC | CAN | 2022 | 2023 | 2024 |
|---|---|---|---|---|
| MH | 8472592 | $515,387 | $515,387 | $544,036 |

Recommended future year total cost support, subject to the availability of funds and satisfactory progress of the project

**NIH Administrative Data:**
**PCC**: 9A-ASPT / **OC**: 41025 / **Released**: Chia, Jackie 05/09/2022
**Award Processed:** 05/10/2022 12:18:19 AM

#### SECTION II – PAYMENT/HOTLINE INFORMATION – 5R01MH122308-03

For payment and HHS Office of Inspector General Hotline information, see the NIH Home Page at http://grants.nih.gov/grants/policy/awardconditions.htm

#### SECTION III – STANDARD TERMS AND CONDITIONS – 5R01MH122308-03

This award is based on the application submitted to, and as approved by, NIH on the above-titled project and is subject to the terms and conditions incorporated either directly or by reference in the following:

a. The grant program legislation and program regulation cited in this Notice of Award.
b. Conditions on activities and expenditure of funds in other statutory requirements, such as those included in appropriations acts.
c. 45 CFR Part 75.
d. National Policy Requirements and all other requirements described in the NIH Grants Policy

Statement, including addenda in effect as of the beginning date of the budget period.

e. Federal Award Performance Goals: As required by the periodic report in the RPPR or in the final progress report when applicable.
f. This award notice, INCLUDING THE TERMS AND CONDITIONS CITED BELOW.

(See NIH Home Page at http://grants.nih.gov/grants/policy/awardconditions.htm for certain references cited above.)

**Research and Development (R&D):** All awards issued by the National Institutes of Health (NIH) meet the definition of "Research and Development" at 45 CFR Part§ 75.2**.** As such, auditees should identify NIH awards as part of the R&D cluster on the Schedule of Expenditures of Federal Awards (SEFA). The auditor should test NIH awards for compliance as instructed in Part V, Clusters of Programs. NIH recognizes that some awards may have another classification for purposes of indirect costs. The auditor is not required to report the disconnect (i.e., the award is classified as R&D for Federal Audit Requirement purposes but non-research for indirect cost rate purposes), unless the auditee is charging indirect costs at a rate other than the rate(s) specified in the award document(s).

This institution is a signatory to the Federal Demonstration Partnership (FDP) Phase VII Agreement which requires active institutional participation in new or ongoing FDP demonstrations and pilots.

An unobligated balance may be carried over into the next budget period without Grants Management Officer prior approval.

This grant is subject to Streamlined Noncompeting Award Procedures (SNAP).

This award is subject to the requirements of 2 CFR Part 25 for institutions to obtain a unique entity identifier (UEI) and maintain an active registration in the System for Award Management (SAM). Should a consortium/subaward be issued under this award, a UEI requirement must be included. See http://grants.nih.gov/grants/policy/awardconditions.htm for the full NIH award term implementing this requirement and other additional information.

This award has been assigned the Federal Award Identification Number (FAIN) R01MH122308. Recipients must document the assigned FAIN on each consortium/subaward issued under this award.

Based on the project period start date of this project, this award is likely subject to the Transparency Act subaward and executive compensation reporting requirement of 2 CFR Part 170. There are conditions that may exclude this award; see http://grants.nih.gov/grants/policy/awardconditions.htm for additional award applicability information.

In accordance with P.L. 110-161, compliance with the NIH Public Access Policy is now mandatory. For more information, see NOT-OD-08-033 and the Public Access website: http://publicaccess.nih.gov/.

This award provides support for one or more clinical trials. By law (Title VIII, Section 801 of Public Law 110-85), the "responsible party" must register "applicable clinical trials" on the ClinicalTrials.gov Protocol Registration System Information Website. NIH encourages registration of all trials whether required under the law or not. For more information, see http://grants.nih.gov/ClinicalTrials_fdaaa/

In accordance with the regulatory requirements provided at 45 CFR 75.113 and Appendix XII to 45 CFR Part 75, recipients that have currently active Federal grants, cooperative agreements, and procurement contracts with cumulative total value greater than $10,000,000 must report and maintain information in the System for Award Management (SAM) about civil, criminal, and administrative proceedings in connection with the award or performance of a Federal award that reached final disposition within the most recent five-year period. The recipient must also make semiannual disclosures regarding such proceedings. Proceedings information will be made publicly available in the designated integrity and performance system (currently the Federal Awardee Performance and Integrity Information System (FAPIIS)). Full reporting requirements and procedures are found in Appendix XII to 45 CFR Part 75. This term does not apply to NIH fellowships.

**Treatment of Program Income:**

Version: 5 - 12/16/2021 7:39 PM | Generated on: 5/10/2022 12:18 AM

**SECTION IV – MH SPECIFIC AWARD CONDITIONS – 5R01MH122308-03**

Clinical Trial Indicator: Yes
This award supports one or more NIH-defined Clinical Trials. See the NIH Grants Policy Statement Section 1.2 for NIH definition of Clinical Trial.

**AWARD NOTICE**:
This award has been made in response to the application submitted under the Funding Opportunity Announcement PA19-042 which can be referenced at: https://grants.nih.gov/grants/guide/pa-files/PA-19-042.html

**INFORMATION:**
This grant is awarded with the understanding that project delays and challenges may occur due to COVID-19. It is NIMH's intention to ensure the ultimate success of each project: to that end, we will work with recipients on a case-by-case basis to identify flexibilities and find solutions. You are encouraged to refer to the NIH Guide (https://grants.nih.gov/grants/guide/notice-files/NOT-OD-20-086.html, and updates referenced therein) and to the regularly updated Frequently Asked Questions (https://grants.nih.gov/faqs#/covid-19.htm) for guidance on delays in research progress, delays in financial and RPPR reporting, costs, and other relevant issues, and contact your grants specialist and/or program officer with questions.

The NIH is concerned about the safety and welfare of human subject participants and research staff and has issued Guidance for NIH-funded Clinical Trials and Human Subjects Studies Affected by COVID-19 . For the purposes of RMR reporting, the NIMH requests that study teams continue to enter their anticipated recruitment milestones and actual recruitment numbers and succinctly indicate in the comments box if and how COVID-19 has impacted study recruitment efforts. NIMH staff will continue to monitor and work with you as additional guidance and information becomes available.

**CONSORTIUM / CONTRACTUAL COSTS**:
This award includes funds for consortium activity with Parnters in Hope (Malawi), Charles University (Czech Republic) and Medical University of South Carolina. Each consortium is to be established and administered in accordance with the NIH Grants Policy Statement (http://grants.nih.gov/grants/policy/nihgps/index.htm). No foreign performance site may be added to this project without the written prior approval of the National Institute of Mental Health.

**PARTICIPANT RECRUITMENT - MILESTONES**:
Future NIMH support for this study is contingent upon adequate participant recruitment based on projected milestones as approved in the Recruitment Milestone Reporting system (RMR) on 6/15/2021. It is expected that **580** of the **1100** total projected participants will be recruited by 4/1/2023. This tri-yearly recruitment report should be submitted electronically to NIMH after each milestone period of April 1, August 1 and December 1 at: http://wwwapps.nimh.nih.gov/rmr/displayHome.action. In the event that actual recruitment falls significantly below projected milestones, NIMH may consider withholding future support and/or negotiating an orderly phase-out of this study. Information regarding the NIMH Policy for the Recruitment of Participants in Clinical Research is available at: https://grants.nih.gov/grants/guide/notice-files/NOT-MH-19-027.html.

**GCP TRAINING**:
NIH expects that all NIH-funded investigators and staff who are involved in the conduct, oversight, or management of clinical trials should be trained in Good Clinical Practice (GCP), consistent with principles of the International Conference on Harmonisation (ICH) as stated in NOT-OD-16-148.

**CLINICAL TRIAL DISSEMINATION PLAN**:
The clinical trial(s) supported by this award is subject to the plan within the competing application dated 9/9/19 submitted to NIH and the NIH policy on Dissemination of NIH-Funded Clinical Trial Information. The plan states that the clinical trial(s) funded by this award will be registered in ClinicalTrials.gov not later than 21 calendar days after enrollment of the first participant and

primary summary results reported in ClinicalTrials.gov, not later than one year after the completion date. The reporting of summary results is required by this term of award even if the primary completion date occurs after the period of performance.

**CLINICAL TRIAL REQUIREMENTS**:
This award is subject to additional certification requirements with each submission of the Annual, Interim, and Final Research Performance Progress Report (RPPR). The recipient must agree to the following annual certification when submitting each RPPR. By submitting the RPPR, the AOR signifies compliance, as follows:
In submitting this RPPR, the SO (or PD/PI with delegated authority), certifies to the best of his/her knowledge that, for all clinical trials funded under this NIH award, the recipient and all investigators conducting NIH-funded clinical trials are in compliance with the recipient's plan addressing compliance with the NIH Policy on Dissemination of NIH-Funded Clinical Trial Information. Any clinical trial funded in whole or in part under this award has been registered in ClinicalTrials.gov or will be registered not later than 21 calendar days after enrollment of the first participant. Summary results have been submitted to ClinicalTrials.gov or will be submitted not later than one year after the completion date, even if the completion date occurs after the period of performance.

**DATA AND SAFETY MONITORING:**
This grant is expected to involve a Data and Safety Monitoring Board (DSMB). The recipient and Principal Investigator(s) should review the NIMH Policy Governing Independent Safety Monitors and Independent Data and Safety Monitoring Boards cited in NOT-MH-19-027. Each RPPR submitted should include copies of all DSMB reports in RPPR Section G.1 since the last annual award. The recipient is also reminded of required reporting pursuant to NIHGPS 4.1.15.3 and the NIMH Reportable Events Policy cited in NOT-MH-19-027, as appropriate.

**SPREADSHEET SUMMARY**
**AWARD NUMBER:** 5R01MH122308-03

**INSTITUTION:** UNIVERSITY OF CALIFORNIA LOS ANGELES

| Budget | Year 3 | Year 4 | Year 5 |
|---|---|---|---|
| Salaries and Wages | $91,521 | $91,521 | $91,521 |
| Fringe Benefits | $25,255 | $25,255 | $25,255 |
| Personnel Costs (Subtotal) | $116,776 | $116,776 | $116,776 |
| Materials & Supplies | $1,800 | $1,800 | $1,800 |
| Travel | $9,270 | $9,270 | $8,505 |
| Other | $325 | $325 | $325 |
| Subawards/Consortium/Contractual Costs | $306,812 | $306,812 | $330,701 |
| Publication Costs | $3,600 | $3,600 | $3,600 |
| TOTAL FEDERAL DC | $438,583 | $438,583 | $461,707 |
| TOTAL FEDERAL F&A | $76,804 | $76,804 | $82,329 |
| TOTAL COST | $515,387 | $515,387 | $544,036 |

| Facilities and Administrative Costs | Year 3 | Year 4 | Year 5 |
|---|---|---|---|
| F&A Cost Rate 1 | 56% | 56% | 56% |
| F&A Cost Base 1 | $137,150 | $137,150 | $147,016 |
| F&A Costs 1 | $76,804 | $76,804 | $82,329 |
