## Supporting Information S2 for "Engaging Men Through HIV Self-Testing with Differentiated Care to Improve ART Initiation and Viral Suppression among Men in Malawi (ENGAGE): a study protocol for a randomized control trial"

**ABSTRACT**

**Background:** Men in sub-Saharan Africa are less likely than women to use HIV services.^1^ HIV testing strategies have dramatically improved for men through HIV self-testing (HIVST) and other male-focused strategies, but men who are identified as living with HIV are still less likely than women to initiate ART or remain in care, particularly within the first several months after initiation. In Malawi, men represent only 31% of new ART initiates^2^ – and men on ART are more likely than women to be lost to follow up (LTFU) at all time intervals of care than women.^3^ Men’s absence from care is concerning not only for their own health, but also for the health of girls and young women who continue to be infected at unacceptably high rates.^4^ Offering ART at home or other locations convenient for men (i.e., home-based ART) for a brief period of time may help men overcome barriers related to facility-based treatment, develop coping strategies for ART engagement, and better engage in facility-based services over the long-term. In this study, we will assess the impact of hbART by comparing two arms:

*Facility-Based ART (fbART arm)*: community-based male-specific counseling followed by linkage to a local health facility for ART initiation and continuation.

*Home-Based ART (hbART arm):* community-based male-specific counseling followed by home-based ART initiation (or at any location that is convenient for participants) and home-based ART continuation for a 3-month period, followed by linkage to a local health facility for further ART continuation.

**Objective:** Our primary objective is to compare the impact of home-based ART against standard of care for ART initiation and retention among men identified as HIV-positive through HIVST in Malawi. Our specific Aims are:

**Aim 1**. Test the effectiveness of hbART versus fbART on ART initiation and 6-month viral suppression among men living with HIV.

**Aim 2.** Identify predictors of ART initiation and 6-month viral suppression in the hbART arm

**Aim 3.** Determine the cost-effectiveness and scalability of hbART at a national level.

**Methods:** We will perform an individually randomized control trial with 700 HIV-positive men who have not yet initiated ART and a subset of 400 of their female partners. Men will be individually randomized 1:1 to one of the two intervention arms described above and will be enrolled in the study for 18-months or until 12-month retention (secondary outcome) can be measured, whichever comes first. The study will be performed at 10 health facilities supported by Partners in Hope (PIH). Data collection will include baseline and follow-up surveys at 2-, 4-, and 7-months, as well as medical charter reviews for men at 2-, 4-, 7-, and 13-months after study enrollment. Qualitative interviews will be conducted with a subset of men and women to understand perceptions of the intervention and experiences with ART engagement, and cost data from a provider perspective will be collected for a cost analysis of each arm.

**Anticipated results**: Findings will establish the effectiveness of home-based ART among men living with HIV who have not yet engaged in treatment, and can directly inform HIV programs throughout the region. Findings will also help us assess if short-term home-based ART is sufficient to engage men in long-term facility-based care, or if additional, more resource-intensive services are needed, such as major changes to the structure of facility-based ART.

### 1. SPECIFIC AIMS

We propose to test a novel strategy to improve ART services targeting HIV-positive men who are not yet engaged in care in Malawi. Men in sub-Saharan Africa are less likely than women to use HIV services.^1^ Men’s absence from care is especially concerning for men who have an HIV-positive female partner (i.e., index client), as they have double the risk of seroconversion compared to other men.^2^

Innovative ART initiation strategies are urgently needed for men. Current standard of care in Malawi is to provide assisted linkage and peer navigation to anyone who tests HIV-positive. However, assisted linkage alone does not addressed known barriers to men’s engagement in ART services. Our formative work found that men avoided ART services due to (1) fear of unwanted disclosure and stigma, (2) time/cost required to access care, (3) poor knowledge about the benefits of early ART initiation, and (4) feeling healthy at the time of testing, therefore there was little perceived motivation or benefit to starting treatment.

In this study, we propose to test a differentiated model of care (DMOC) intervention for ART initiation and 6-month retention that includes (a) home-based ART initiation and continuation for 3-months (hbART), and (b) assisted linkage to facility ART at the end of three months. The intervention will be compared to standard facility-based ART (fbART). The intervention will include: (visit1) home-based ART initiation + client-centered counseling; (visit2 & 3) home-based ART + client-centered counseling; (visit4) assisted facility linkage + peer navigation to integrate into fbART. We hypothesize that hbART will largely remove barriers to ART initiation, and provide coping strategies men need to overcome barriers related to facility-based retention. Specific Aims are:

**Aim 1. Test the effectiveness of hbART versus fbART on ART initiation and 6-month viral suppression among men living with HIV.** We will conduct a randomized trial with two arms: (1) hbART; and (2) fbART. Men will be recruited from medical chart reviews and from female ART clients who may have partners who participated in index HIV testing and have a male partner living with HIV. Men will be recruited, screened, and enrolled at their homes or other locations convenient for them.

***Hypothesis 1.1.*** ART initiation will be at least 40% greater in the hbART arm compared to fbART arm.

***Hypothesis 1.2.*** Six-month viral suppression will be at least 20% greater in the hbART arm compared to fbART, using intention-to-treat analyses.

**Aim 2. Identify predictors of ART initiation and 6-month viral suppression in the hbART arm.** We will conduct baseline surveys with men and a sub-set of their partners (female ART clients with male partners who are living with HIV) to capture sociodemographic and biomedical predictors of care uptake across the treatment cascade (ART initiation, 6-month viral suppression). We will conduct qualitative interviews with a subset of both men and female partners to assess in-depth reasons men succeed or fail to initiate ART and reach viral suppression.

***Hypothesis 2.1.*** Primary predictors of ART initiation and 6-month viral suppression will include: perceptions of ART services (male-friendly), couple characteristics (joint-decision making), and perceived need (ART knowledge and perceptions).

**Aim 3. Determine the cost-effectiveness and scalability of hbART at a national level.** We will assess cost-effectiveness using a facility perspective.

***Hypothesis 3.1.*** hbART will have an incremental cost-effectiveness ratio (derived from comparison of the cost per outcome metrics between the intervention and control arms) that is deemed to be of policy significance based on comparisons to cost-effectiveness thresholds.

### 2.  LITERATURE REVIEW

***Background***

Men in sub-Saharan Africa are less likely than women to use HIV services.^5^ Men’s absence from care is concerning not only for their own health, but also for the health of girls and young women who continue to be infected at unacceptably high rates.^6^ HIV prevention and treatment programs have not traditionally been directed at men. Men are notably absent from international guidelines, national policies, and local HIV interventions. Research shows that women are 322% more likely to be mentioned in international HIV guidelines than men.^7^ In the context of Malawi, national guidelines expect women of reproductive age to attend a health facility 5-17 times per year (equivalent to 19-63 hours),^8^ and 180-472 times in their reproductive lifespan (15-44 years). There are no such expectations for men (see Table 1). The justification for the global attention of HIV program thus far on women and girls is without dispute. Gender inequality is a key driver which impacts women's health and access to HIV services and creates specific vulnerabilities for women to HIV infection.^9^ However, framing HIV as a woman’s concern means we have failed to understand how gender affects and drives the burden of ill health for men, and inadvertently perpetuates the epidemic for young women and girls. Targeted strategies specific to men are urgently needed if we are to engage them in care.

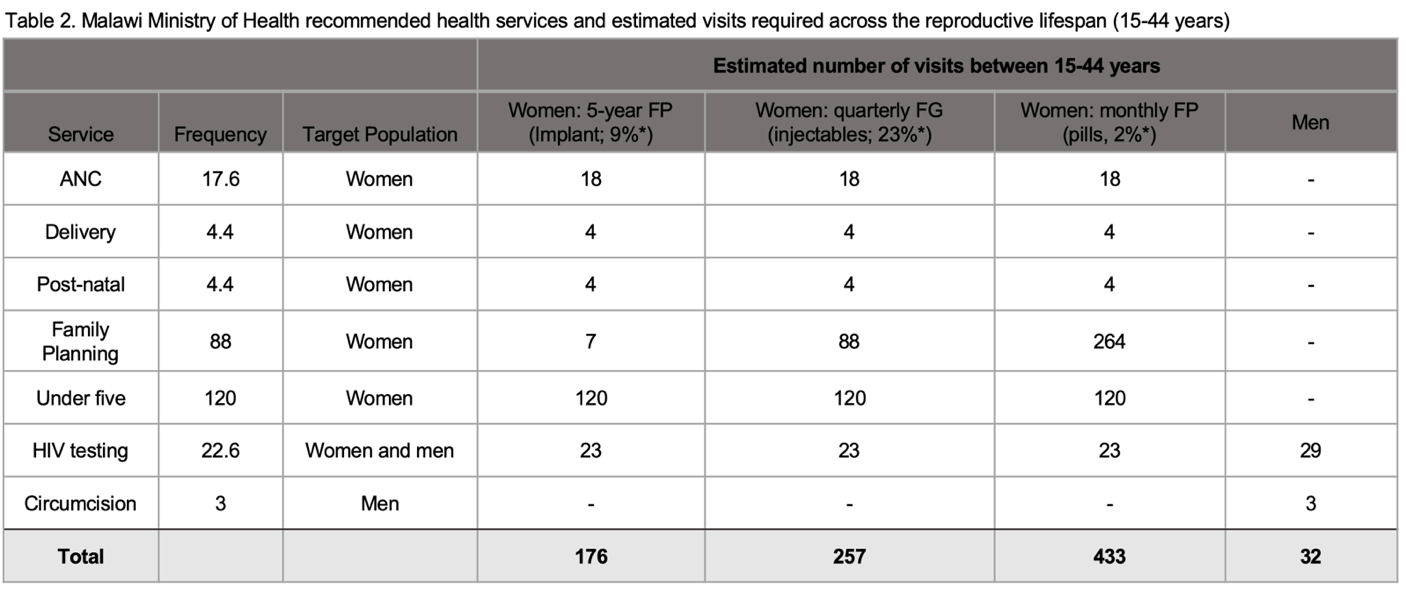
***Table 1:*** *Malawi ministry of health recommended health services and estimated visits required across the reproductive life span (15-44 years)*

Men in sub-Saharan Africa are less likely than women to use HIV services.^1^ HIV testing strategies have dramatically improved for men through HIV self-testing (HIVST) and other male-focused strategies, but men who are identified as living with HIV are still less likely than women to initiate ART or remain in care, particularly within the first several months after initiation. In Malawi, men represent only 31% of new ART initiates^2^ – and men on ART are more likely than women to be lost to follow up (LTFU) at all time intervals of care than women.^3^ Men’s absence from care is concerning not only for their own health, but also for the health of girls and young women who continue to be infected at unacceptably high rates.^4^ Offering ART at home or other locations convenient for men (i.e., home-based ART) for a brief period of time may help men overcome barriers related to facility-based treatment, develop coping strategies for ART engagement, and better engage in facility-based services over the long-term.

Two overarching barriers keep HIV-positive men from accessing ART services: (1) lack of male-friendly services;^10-12^ and (2) harmful gender norms.^13-15^ Male friendly services are private and convenient (requiring minimal time), and offered by health workers who understand the unique needs of men.^16^ In addition, men are often unfamiliar with the health system, and are unsure how to navigate facility-based services. Gender norms that prioritize men as strong and self-reliant perpetuate fear of unwanted disclosure and stigma, and discourage men’s engagement in ART.^13,16^ Our research in Malawi found similar barriers to ART initiation for men who tested HIV-positive: men avoided ART services due to (1) fear of unwanted disclosure and stigma due to lack of privacy; (2) time/cost required to access care; (3) poor knowledge about the benefits of early ART initiation; and (4) beliefs that require men be strong, in control, and focused on short-term benefits such as daily financial earnings and respect from their male friends. Index HIVST must be combined with innovative ART interventions that address these barriers.

***Evidence based for interventions that increase ART initiation***

We have conducted a thorough search of the literature and have identified several intervention strategies that may increase ART initiation among men who use HIVST: (1) client-centered counseling and motivational interviewing; and (2) home-based ART.

*Client-centered counseling* is becoming widely recognized as a key strategy to help clients navigate barriers to the desired outcome by building client’s self-efficacy, identifying internal motivation for the desired behavior, and establishing strategies and short- and long-term goals needed to reach a desired outcome.^17,18^ Motivational interviewing is seen as particularly effective when clients need to make difficult decisions and overcome multi-level barriers to behavior change.^19,20^ The strategy has been used to improve ART adherence^17,19^ and reduce sexual risk behavior.^11^

In contrast, traditional counselling efforts are largely informational and directive, whereby health care workers deliver a pre-determined counseling package that is not responsive to a client’s individual situation.^10,12^ Such methods have been proven largely ineffective,^13^ particularly with hard-to-reach populations such as men.^15^ Client-centered counseling tailored specifically to men differs from traditional strategies by adopting a client-centered approach is based on collaboration, evocation and respect for autonomy. We hypothesize that these counseling techniques will encourage HIV status acceptance and disclosure, promote health seeking behavior, provide coping strategies men need to overcome barriers related to facility-based care, and ultimately, facilitate ART initiation.

*Home-based ART initiation* has improved ART initiation across the region. A systematic review found that home-based ART is associated with ART retention, decreased mortality,^21^ and in some cases, reduced stigma and increased privacy.^22,23^ We conducted one of the only studies to examine home-based ART initiation within a community HIVST distribution strategy (Co-I: Choko). We found that home-based ART initiation alongside home-based HIVST significantly increased ART initiation as compared to standard facility-based initiation (RR 2.94; p-value<0.001).^24^

Home-based ART may be particularly attractive to harder-to-reach men because it reduces client time required to access services and provides an easy, opt-out entry point for men who otherwise may have never engaged with the health care system, or know how to navigate complicated, busy health facilities. Home-based ART has been associated with a three-fold reduction in financial costs to clients.^25^ Further, home-based ART facilitates client-centered, one-on-one care that is often not feasible in busy clinic settings.

Finally, increased privacy and decreased wait-times are essential if men are to engage in HIV services. As part of a study on new Universal Treatment policies in Malawi, 15 in-depth interviews and 208 surveys were conducted with newly diagnosed HIV-positive men.^26,27^Fear of unwanted disclosure due to limited privacy and a lack of trust in the health facility were the primary barriers to men’s ART initiation. Home-based ART initiation can help address these barriers for ART initiation, and motivational interviewing can help provide men the skills needed to navigate these barriers within the health system in order to promote ART retention. Further, we find in our formative work that the vast majority (>95%) of men who used HIVST in the Index HIVST trial disclosed their HIV status to their female partner,^17^ meaning that home visits (i.e., reminders, peer navigation, motivational interviewing, or home-based ART) will not increase risk of unwanted disclosure to one’s sexual partner. Table 2 outlines how the proposed interventions will address barriers to ART initiaiton identified in the literature.

**Table 2.** Barriers and associated intervention components

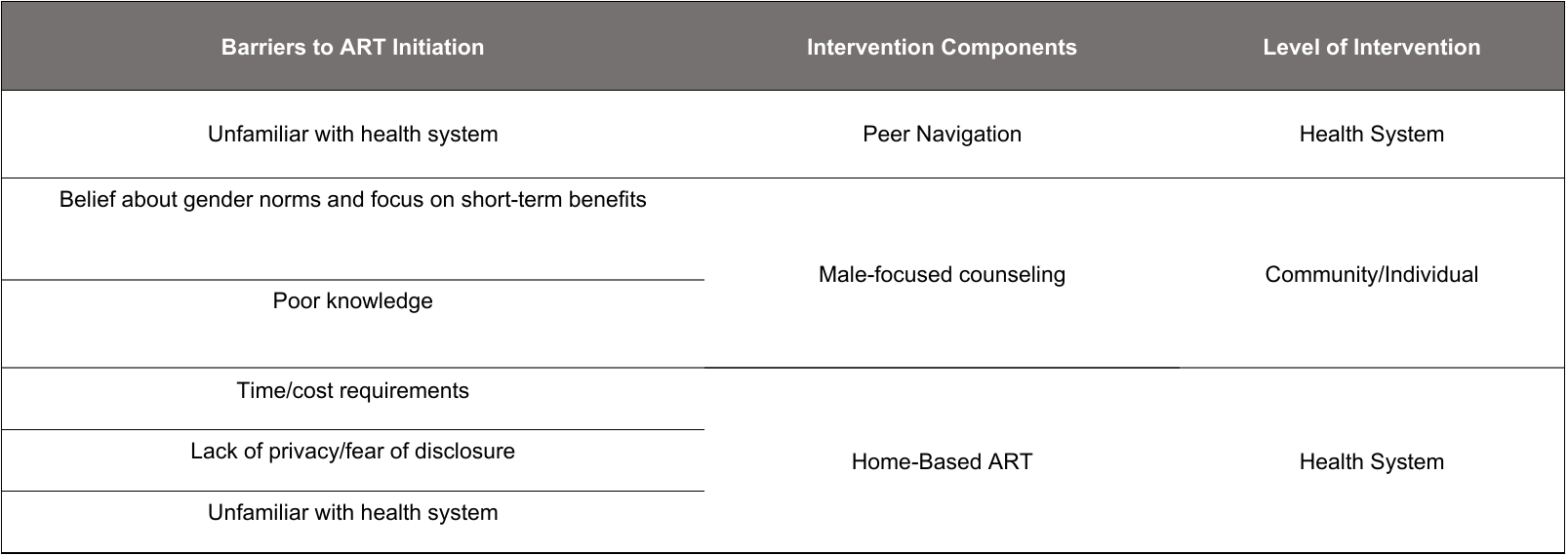

### 3. METHODOLOGY

We will perform an individually randomized control trial with 700 HIV-positive men who have not yet initiated ART and a subset of 400 of their female partners to improve ART initiation, retention and viral suppression among men who test HIV-positive but have not yet initiated ART. Men will be individually randomized 1:1 to one of two intervention arms: fbART or hbART. Our primary outcome of interest is 6-month viral suppression, but we will also collect data on ART initiation, 6-month retention, and 12-month retention. Data collection will include baseline and follow-up surveys at 2-, 4-, and 7-months, as well as medical charter reviews for men at 2-, 4-, 7-, and 13-months after study enrollment. Qualitative interviews will be conducted with a subset of men and women to understand perceptions of the intervention and experiences with ART engagement, and cost data from a provider perspective will be collected for a cost analysis of each arm. Men will be enrolled in the study for 18-months or until 12-month retention (secondary outcome) can be measured, whichever comes first. Figure 2 depicts how participants move through the study.

**Figure 2:** *Study flow chart*

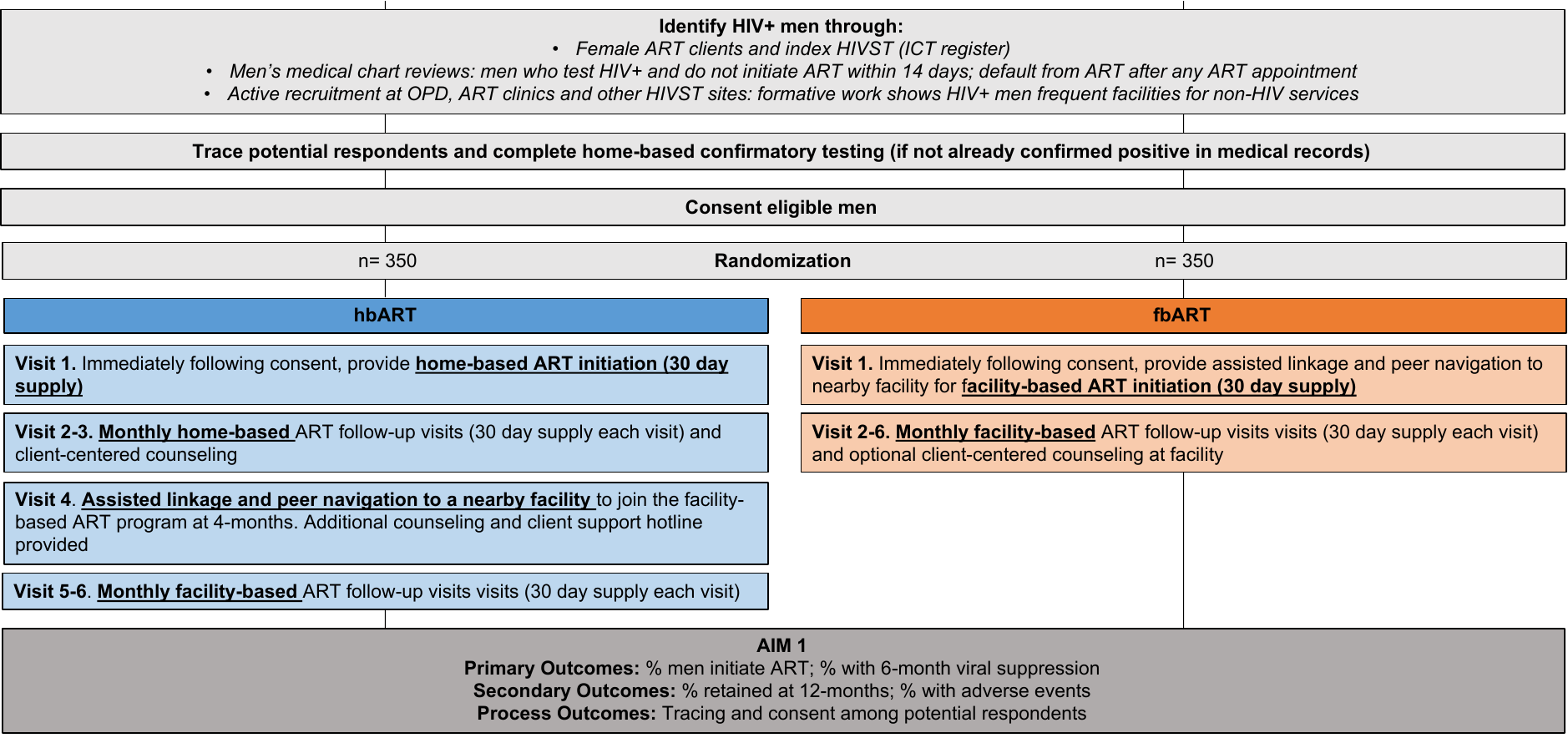

#### 3.1. Place of study

All study activities will take place in 10 Partners in Hope supported facilities (See Table 3 below). These sites were chosen because they are priority districts for the Presidents Emergency Plan for AIDS Relief (PEPFAR) and have the highest HIV prevalence and unmet need of all Partners in Hope supported districts.

***Table 3:*** *Selected study sites*

| **District** | **Facility name** | **ART Cohort Size** |
| --- | --- | --- |
| Chikwawa | St. Montfort Mission Hospital | 4208 |
| Chikwawa | Ngabu Rural Hospital | 4185 |
| Chikwawa | Chikwawa District Hospital | 6377 |
| Dowa | Mponela Rural Hospital | 2239 |
| Lilongwe | Daeyang Luke Hospital | 2113 |
| Lilongwe | Likuni Mission Hospital | 4288 |
| Nkhotakota | Alinafe Community Hospital | 897 |
| Nsanje | Kalemba Community Hospital | 2437 |

#### 3.2. Intervention description

Participating men will be offered an intervention immediately after study enrollment. A description of study arms is below:

**Facility-based ART arm:** Male participants will also be offered Men’s Specific Counselling at male participant’s home, or a nearby location that is preferred by participants, followed with an escort to the facility and “warm handover” with facility navigation (1 home-visit in total). The initial home visit will be scheduled at times convenient for men, including evening and weekend hours. Men’s Specific Counseling includes counseling on the benefits of early ART, strategies for disclosure and positive living, strategies to overcome facility-based barriers to ART services, and addressing harmful gender norms that may discourage men from using care. Counseling will be adaptive to the needs and concerns of male clients.

Male participants who refuse ART initiation will receive 3 additional visits in 14-day intervals to offer counselling and encourage initiation.

**Home-based ART arm:** ART initiation and two follow-up appointments will be given at male participant’s home, or a nearby location that is preferred by participants (3 home-visits in total) - meeting spaces must be within the facility catchment area. Home-based ART appointments will be scheduled at times convenient for men, including evening and weekend hours. Male participants will also be offered Men’s Specific Counselling, as with the fbART arm. Participants will be escorted to the nearest health facility for their third follow-up appointment, and given facility navigation and a “warm handover” to routine facility care. All intervention activities in this Arm will be offered by a trained Male Nurse. Details of specific visits are below:

- **Visit 1:** The study (male) nurse will visit the participant at a time and location convenient for the participant and provide Male Specific Counseling.
  Those who agree to initiate ART will receive the following:
  - Basic general health evaluation- include TB screening, WHO staging, and POC CD4 count (if available at facility). Participants with advanced HIV and any potential complications will be immediately referred and escorted to the facility for facility-based ART initiation.
  - ART initiation with a 30-day supply of first-line ART in Malawi – dolutegravir, tenofovir, and lamivudine as a single tablet
  - 30-day supply of cotrimoxazole, standard of care for all HIV-positive individuals.
  - Schedule time and place for home-based 4-week follow-up ART refill appointment

Male participants who opt for facility-based care instead of home-based care will be referred and escorted to facility, and given a “warm handover”, including facility navigation.

Male participants who refuse ART initiation will receive 3 additional visits in 14-day intervals to offer counselling and encourage initiation.

- **Visit 2 & 3:** The study (male) nurse will visit the participant at 1- and 2- month intervals, aligning with their 1st and 2nd ART refill appointments. Those who agree to initiate ART will receive the following:
  - Basic general health evaluation- include TB screening, WHO staging, and POC CD4 count (if available at facility) -- participants with advanced HIV and any potential complications will be immediately referred and escorted to the facility for facility-based ART initiation.
  - ART initiation with a 30-day supply of first-line ART in Malawi – dolutegravir, tenofovir, and lamivudine as a single tablet
  - 30-day supply of cotrimoxazole, standard of care for all HIV-positive individuals.
  - Schedule time and place for next monthly appointment.
- **Visit 4:** The study (male) nurse will escort the participant to the facility and provide a “warm handover” with facility navigation and male-specific counseling. The 3rd ART refill and all subsequent ART services will be provided at the health facility.

Specific methodology for each Objective is described below:

#### 3.3. Aim 1 & Aim 2

**Aim 1.** Test the effectiveness of hbART versus fbART on ART initiation and 6-month viral suppression among men living with HIV.

**Aim 2.** Identify predictors of ART initiation and 6-month viral suppression in the hbART arm

##### 3.3.1. Study design

We will conduct an unblinded, individually randomized controlled trial at 10 high-burden facilities in Malawi, randomizing individual men 1:1 to fbART and hbART arms.

We chose individual-level randomization for several reasons. First, individual-level randomization allows us to control for potential spurious relationships between the primary outcome (6-month retention) and the intervention that may be due to unobserved facility- and district-level factors. Other studies show high variation in ART outcomes by site, even within the same district, due to facility staff, site infrastructure, and unmeasurable factors.^56^ Second, programmatic strategies for HIV services are rapidly changing and are likely to vary by site. We will use individual-level randomization to ensure that all three arms are exposed to similar external activities.

##### 3.3.2. Target population

We will enroll 700 HIV-positive men who are not currently engaged in ART services. We will also enroll 400 female partners of HIV-positive men who are enrolled in the ART program at participating facilities and self-report that their male partner is HIV-positive and not currently engaged in ART.

###### ***Eligibility Criteria***

*Female Partner*

While the study’s primary focus is men, a subset of female partners will be enrolled in order to help recruit men and to understand female partners’ perception of of the interventions and any unintended outcomes or adverse events for female partners that may be associated with the interventions.

Inclusion criteria:

All of the following criteria must be met in order for men to be eligible for the study. All criteria for female partners is based on self-report from the female partner:

- ≥15 years of age
- Male partner ≥15 years of age
- No reported interpersonal violence (IPV) as defined by WHO with the above male partner in the past 12-months
- Male partner ever tested HIV-positive
- Male partner not currently engaged in ART services, defined as:
  - Tested HIV-positive ≥14 days and not on ART ≥14 days after testing HIV-positive;
  - ≥14 days late for the first four-week follow up appointment; or
  - Initiated ART but ≥60 days late for last ART appointment;
- Male partner living inside the facility catchment area (defined as any area that HCWs from the study facility routinely visit for tracing purposes)

Exclusion criteria:

Individuals will be excluded if any of the following exclusion criteria are present:

- <15 years of age
- Male partner <15 years of age
- Reported interpersonal violence (IPV) as defined by WHO with the above male partner in the past 12-months
- Male partner never tested HIV positive
- Male partner tested HIV-positive <14 days ago
- Male partner currently engaged in ART services, defined as:
  - Initiated ART
  - ≤14 days late for the first four-week follow up appointment
  - Initiated ART and ≤60 days late for last ART appointment
- Male partner living outside the facility catchment area (defined as any area that HCWs from the study facility routinely visit for tracing purposes)

###### *Men Living with HIV*

###### Men will be enrolled as the primary recipient of the interventions and will account for the primary outcomes of interest. All of the following criteria must be met in order for men to be eligible for the study:

Inclusion criteria:

All of the following criteria must be met in order for men to be eligible for the study:

- ≥15 years of age
- Tested HIV positive using Ministry of Health standard algorithm (Determine + Unigold)
- Not currently engaged in ART services, defined as:
  - Tested HIV-positive ≥14 days and not on ART ≥14 days after testing HIV-positive;
  - ≥14 days late for the first four-week follow up appointment; or
  - Initiated ART but ≥60 days late for last ART appointment;
- Has not taken ART in the past 7-days, as indicated by a point of care (POC) urine assay
- Living inside the facility catchment area (defined as any area that HCWs from the study facility routinely visit for tracing purposes)

Exclusion criteria:

Individuals will be excluded if any of the following exclusion criteria are present:

- <15 years of age
- Never tested HIV positive using Ministry of Health standard algorithm (Determine + Unigold)
- Tested HIV-positive <14 days ago
- Currently engaged in ART services, defined as:
  - Initiated ART
  - ≤14 days late for the first four-week follow up appointment
  - Initiated ART and ≤60 days late for last ART appointment
- Has taken ART in the past 7-days, as indicated by a point of care (POC) urine assay
- Living outside the facility catchment area (defined as any area that HCWs from the study facility routinely visit for tracing purposes)

##### 3.3.3. Sampling techniques and enrollment

Strategies to identify potential participants will be embedded within routine health facility activities. We will prioritize enrollment from index HIVST strategies, but will also recruit from other routine HIV testing and treatment strategies, as well as in person recruitment at OPD and ART clinics (see Figure 3). Trained male study research assistants will conduct all register reviews and recruitment of potential participants. Potential participants will be documented in a study register (Potential Participant Register) using unique identifiers.

***Figure 3:*** *Sampling techniques*

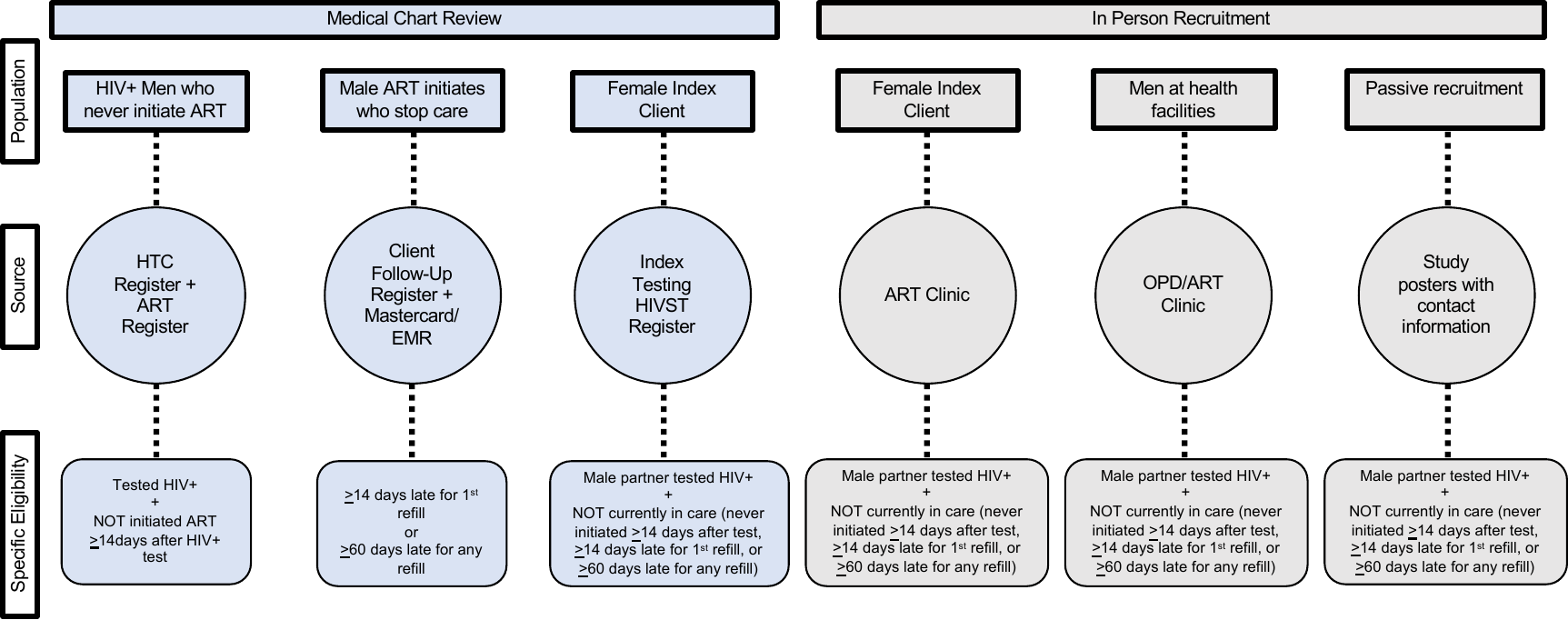

##### 3.3.4. Study Activities

All data collection and intervention activities will be conducted by trained study staff and will be conducted in the local language, Chichewa. Any written material intended for participants will be provided in the local language and using simple language (4^th^ grade reading level). All data will be documented on an encrypted, password protected tablet using a secure data collection software called SurveyCTO and immediately sent to a secure server. Study registers will be used to track daily activities in study sites and will be kept in secure, locked cabinets at participating health facilities – only study staff will have access to study data collection or tracking materials.

Study staff will be comprised of two teams responsible for different components of study activities: 1) the Data Team, which are trained male Research Assistants who are responsible for all study enrollment and data collection activities; and 2) Implementation Team, which are trained health care workers who are responsible for all intervention implementation.

***Female Participants***

Female partners will be engaged by the Data Team. Women will not be contacted by the Implementation Team since no intervention strategies are targeting female participants. See Figure 4 for a description of all study activities to be completed with female participants.

**Figure 4.** Study activities for female participants throughout the study period

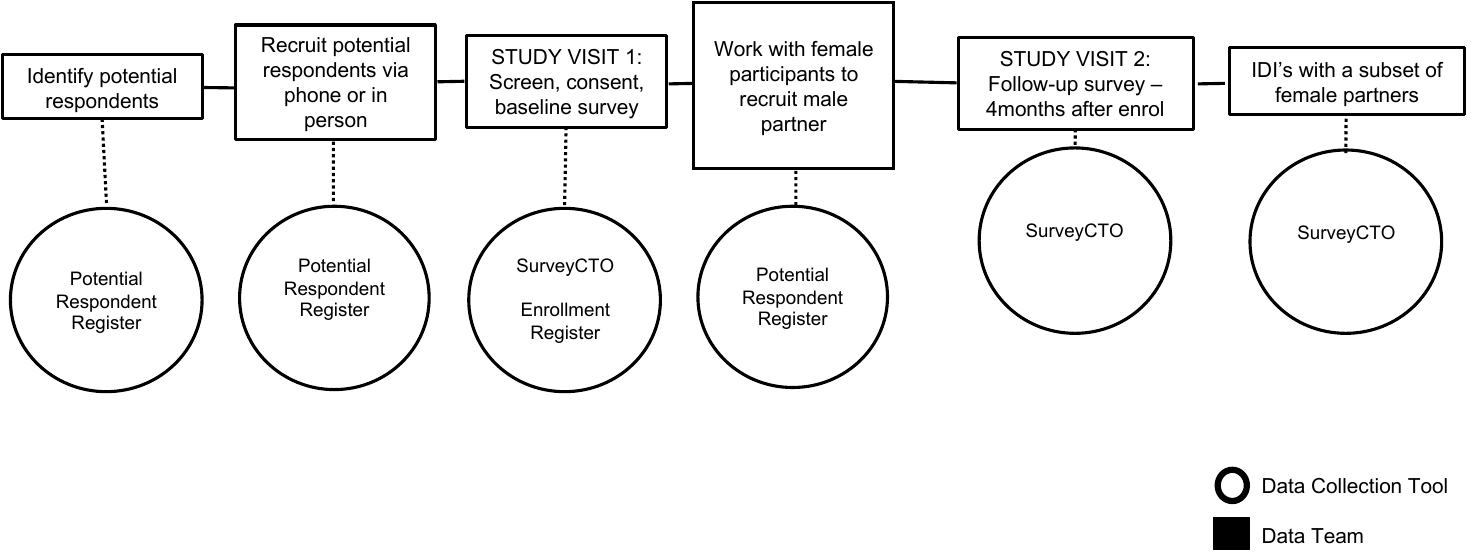

*Female ART client recruitment*

The Data Team will review the routine Index Testing Register on a regular basis to identify female ART clients who reported HIV-positive male partners through routine Index testing procedures. Female ART clients identified as potentially eligible for the study will be contact by the Data Team to assess interest and eligibility for the study. Females will first be contacted via phone, using a phone recruitment script, ensuring that no information about the study is given until the individual’s identity is confirmed (see Appendix A). If after three attempts they cannot be contacted via phone, study staff will trace the potential female participant at their home, using information provided in routine facility medical records. If after three attempts they cannot be contacted in person, the individual will be documented as “unreachable”.

*Female ART client enrollment*

Female partners interested in the study will meet a Research Assistant in person at a place and location that is convenient for them. They will provide oral consent to participate in study screening and be screened for eligibility for the study (Appendix B). Study staff will conduct all consent and screening activities. If eligible and willing to participant, written informed consent will be obtained (Appendix C). Female partners will be enrolled even if they do not think their male partner will agree to participate in the study, but they must be willing to invite their male partner to participate in the study.

Upon enrollment in the study, female ART clients will work with study staff to make a plan for inviting male partners to enroll in the study. Study staff will work with the female ART client to establish a recruitment plan that is acceptable and feasible for the women. This may include women referring male partners to the study staff, study staff actively recruiting male partners based on information provided by the female client, or a joint approach where the study staff approaches male partners with the female ART client. Each female partner will be allowed to choose the recruitment strategy that best fits her individual situation and the needs of her partner.

*Baseline*

The Data Team will administer baseline surveys with women immediately following enrollment. Surveys will last approximately 40 minutes and will collect data on male and female demographics, sexual partnerships and couple dynamics, and men’s history with health services, and HIV services specifically. (Appendix E)

*Follow-up*

The Data Team will administer one follow-up survey with women 4-months after enrollment. Surveys will last approximately 40 minutes and will collect data on changes in male and female demographics, changes in sexual partnerships and couple dynamics, perceptions of the intervention, and any unintended or adverse events (i.e., IPV, end of the relationship, or unwanted disclosure) associated with intervention procedures. (Appendix H).

*In-Depth Interviews*

A subset of women enrolled in the overarching trial will be randomly selected to complete an in-depth interview. Table 4 describes in-depth interview participants and justification for their inclusion. In-depth interview guides for men and women are developed based on existing literature and our extensive experience conducting in-depth interviews with this population (Appendix J and K). Female partners will provide important insight into the circumstances of these men, potential strategies to more effectively reach them, and any adverse events associated with interventions.

***Table 4.*** *Description of in-depth interviews with female partners and justification*

| **Participant type** | **Number of interviews** | **Justification** |
| --- | --- | --- |
| Women whose male partners were **not enrolled** in the study | 30 (~ 15 per arm) | To understand the couple dynamics and characteristics of men who were never traced and additional strategies to reaching these men |
| Women whose male partners **did not initiate ART** | 30 (~ 15 per arm) | To understand the couple dynamics and characteristics of men who were loss-to-follow-up, and additional strategies to better engage these men in care |

***Male Participants***

See Figure 5 for a description of all study and intervention activities to be completed with male participants. Men will engage with both the Data Team and Implementation Team on a regular basis, however Study Visits with the Data Team will always be distinct from Implementation Visits in order to reduce reporting bias due to social desirability concerns (the study staff implementing interventions will never be present when outcome or acceptability data is being collected).

**Figure 5.** Study activities for male participants throughout the study period

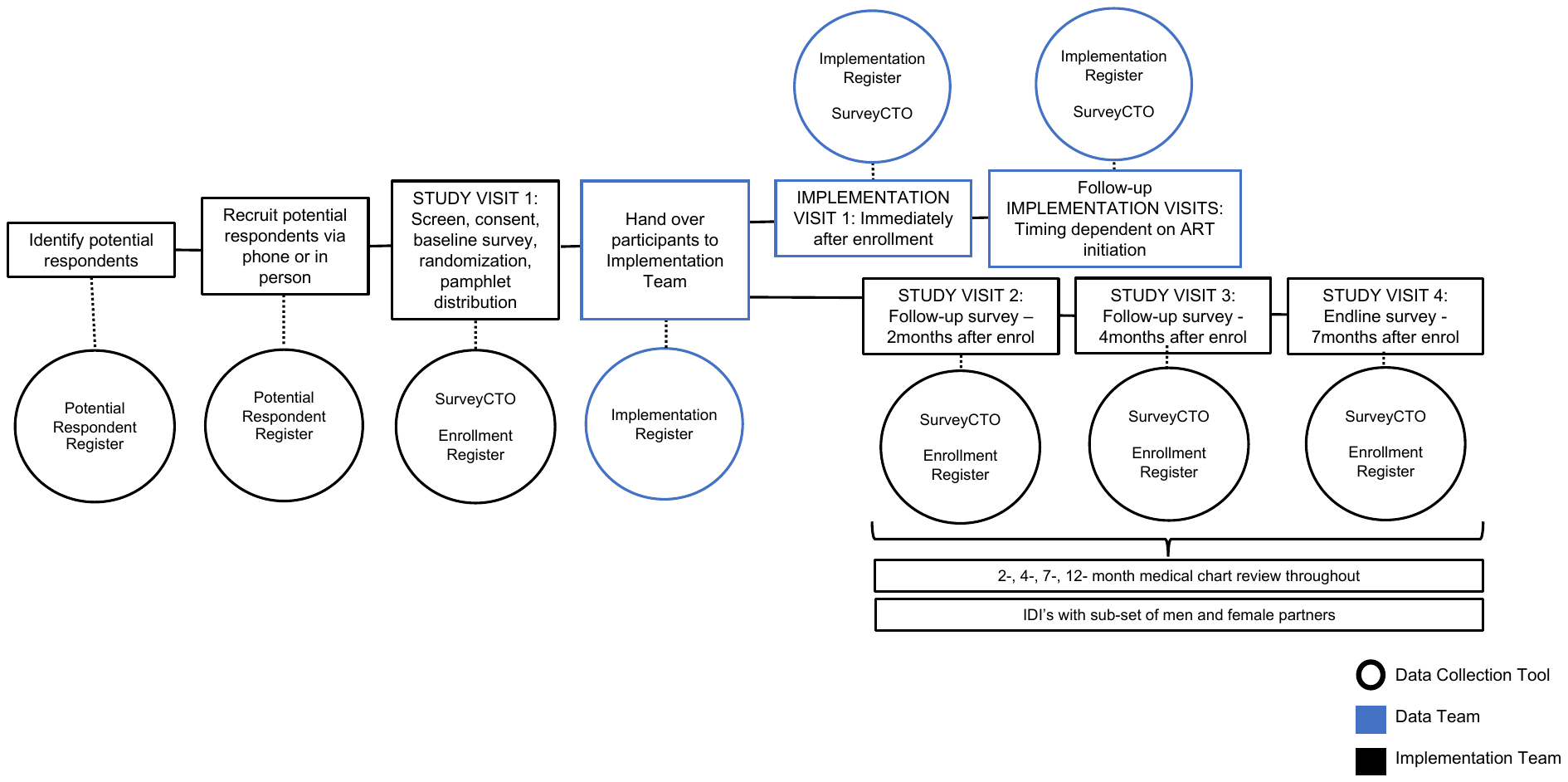

*Male recruitment*

The Data Team will contact potential male participants to assess eligibility and interest in the study. Contact information will come from either routine facility medical records that were used to identify potential participants (refer back to Figure 3) or from enrolled female partners who reported men’s contact information. Men will first be contacted via phone, using a phone recruitment script, ensuring that no information about the study is given until the individual’s identity is confirmed (see Appendix A.2). If after three attempts they cannot be contacted via phone, study staff will trace the potential participant at home, if address information is provided. If after three attempts they cannot be contacted in person, the individual will be documented as “unreachable”.

*Male enrollment*

Men interested in the study will meet study staff in person at a place and location that is convenient for them. During this visit study screening and enrollment, baseline survey, randomization, and introduction to the assigned arm will be conducted (see Figure 6). Men will first provide oral consent to participate in study screening and be screened for eligibility for the study (Appendix B.2).

Potential participants who did not previously initiate ART at participating health facilities and are unable to show proof of confirmatory HIV testing (using Unigold, following Ministry of Health guidelines) via medical records (either personal medical records or facility records) will receive a confirmatory HIV test from a trained HIV Diagnostic Assistant during the eligibility screening process. A confirmatory HIV-positive test must be provided prior to enrollment.

Men who meet all other eligibility criteria will complete a point of care (POC) urine assay in order to have a biomarker measuring exposure to ART in the past 7-days (see UCSF letter of support for urine test strips).^58-60^ Men unable to produce a urine sample will receive a follow-up visit the next day to collect the sample. Biomedical measures to confirm clients are ART naïve are needed for this population since men may be hesitant to disclose current ART use. The assay is processed in 20 minutes and can be conducted at a man’s home or other convenient locations. Those with a reactive urine assay (i.e., had exposure to ART in the past 7-days) are not eligible for the study and will be referred to the local health facility.

Men found eligible and willing to participant, will provide written informed consent (Appendix D). Ineligible men and those who do not consent will be referred to fbART for routine ART.

**Figure 6.** Study Visit 1: Male participant screening, enrollment, baseline, and randomization

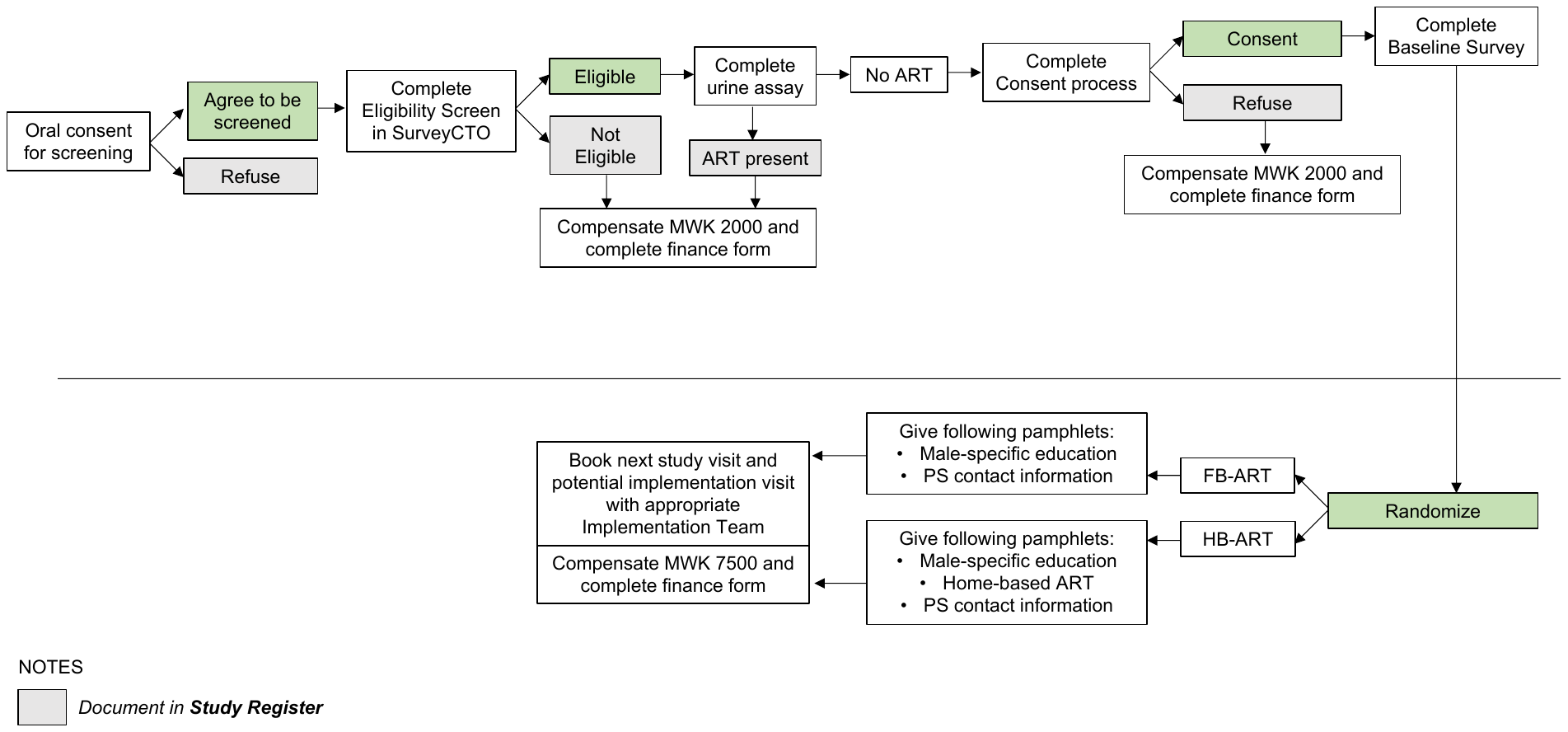

*Baseline*

Research assistants will administer baseline surveys with men immediately following enrollment (before randomization). Surveys will take approximately 40 minutes and will collect data on sociodemographics, sexual partnerships and couple dynamics, and men’s history with health services. (Appendix F)

*Randomization*

Consented men will be randomized 1:1 using an electronic randomization system on tablet devices. Study staff will show randomization results as a picture on a pre-programmed tablet, allowing the participant to view the results themselves in order to maximize transparency and study buy-in.

*Introduction to Arm*

Upon randomization, the Data Team will describe the assigned arm and deliver the following materials.

- **fbART Arm:** Male-specific counseling pamphlet; contact information for the Implementation Team member overseeing implementation of fbART at that site.
- **hbART Arm:** Male-specific counseling pamphlet; information about hbART; contact information for the Implementation Team member overseeing implementation of fbART at that site.

The Data Team will work with each male participant to schedule a tentative meeting time and location for the Implementation Team to visit the participant, and deliver participant information and appointment times to the Implementation Team at the end of each day.

*Intervention Implementation*

The Implementation Team will meet with male participants at the scheduled appointment time or as soon as possible.

For the **fbART Arm**, a Patient Supporter, a lower-cadre staff trained in HIV counseling and mentorship, will be responsible to meet with men at a time and location convenient for men and to provide male-specific counseling, and for those who agree to initiate ART, escort to a local health facility and facility navigation (a “warm handover”). Facility navigation includes orientating men to facility procedures, answer questions that may arise, and working with men to develop a plan to overcome perceived barriers to fbART. Men will receive male-specific counseling during follow-up fbART visits for the first 6-months of care. Men who do not initiate ART after the first visit will be followed up 3-times in roughly 14-day intervals to encourage ART initiation, provide any additional counseling, and answer any questions that arise. These contact points may be via phone or in person and will be conducted by the same Patient Supporter who oversees the fbART Arm.

For the **hbART Arm**, a Study Nurse trained in ART service delivery will be responsible to meet with men at a time and location convenient for them. See Figure 7 for all activities completed during the first Implimentation Visit. During the first visit, the Study Nurse will provide male-specific counseling, and for those who agree to initiate ART, will screen men for eligibility for home-based ART using a validated screening tool to identify individual with WHO Stage 3 or 4 or other opportunistic infections that may require facility-based care. Those who are eligible will be given hbART services. Those who are not eligible will be escorted to a local health facility for fbART and provided facility navigation (a “warm handover”).

Men who do not initiate ART after the first visit will be followed up 3-times in roughly 14-day intervals to encourage ART initiation, provide any additional counseling, and answer any questions that arise. These contact points may be via phone or in person and will be conducted by the same Study Nurse who oversees the hbART Arm.

Men who initiate hbART will receive 2 monthly ART refills at home, along with additional male-specific counseling. At the 3^rd^ ART refill, the Study Nurse will escort men to a local health facility for fbART and provide facility navigation (a “warm handover”), where men will continue receiving fbART for all future months.

#### See “Section 3.2. Intervention description” for further detail on each arm.

Figure 7. HB-ART Implementation Visit 1 Flow Chart: ART Initiation

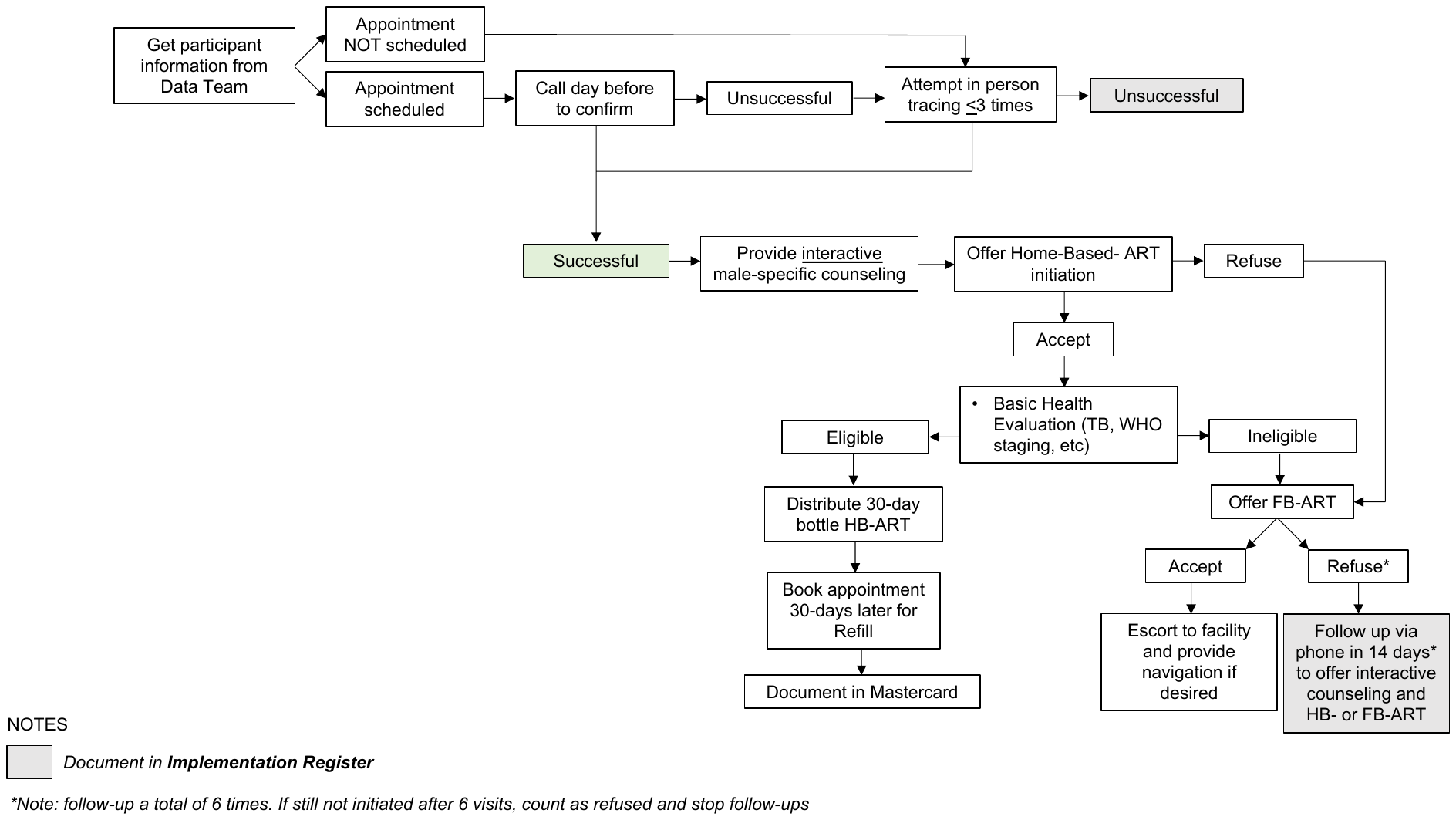

*Follow-up Survey*

Research assistants will administer follow-up surveys and review of health passports with men at 2-, 4-, and 7-months after enrollment in the study. Follow-up surveys will last 40-minutes and will assess exposure to the intervention, acceptability of the intervention, any important changes to sociodemographic factors, and any adverse events (i.e., unwanted status disclosure). (Appendix G)

*In-Depth Interviews*

A subset of men enrolled in the overarching trial will be randomly selected to complete an in-depth interview to be conducted at a time and location that is convenient for the participant. Table 5 describes in-depth interview participants and justification for their inclusion. (Appendix J).

***Table 5.*** *Description of in-depth interview participants and justification*

| **Participant type** | **Number of interviews** | **Justification** |
| --- | --- | --- |
| Male participants who **did not** reach 6-month viral suppression | 30 (~ 15 per arm) | To understand what they liked and did not like about the intervention, why they did not link to ART, and suggestions on how to improve the intervention |
| Male participants who **reached 6-month viral suppression** | 30 (~ 15 per arm) | To understand what they liked and did not like about the intervention, why they linked to ART, and suggestions on how to improve the intervention |

##### 3.3.5. Summary of data collection

- *Baseline and Follow-Up Surveys:* Baseline surveys will be conducted with men enrolled in the trial. Follow-up surveys will be conducted with both men and women enrolled in the study. They will focus on:
- *Health care system*: perceptions regarding the following aspects of ART services (i) privacy and confidentiality; (ii) availability of services; (iii) wait-time and distance to facility; (iv) quality of care and rude behavior from health care providers, using validated measures.
- *Sociodemographics*: (i) age; (ii) household assets; (iii) work; and (iv) substance use
- *Couple characteristics*: (i) relationship type and length; (ii) sexual activity and risk; (iii) frequency of communication; (iv) disclosure; (v) joint decision making using standard measures from Demographic Health Survey (DHS);^28^ (vi) gender norms using validated Gender Equitable Men (GEM) Scale,^29^ and (v) Revised Conflict Tactics Scale.^30^
- *Knowledge/perceptions and biomedical factors:* (i) knowledge about HIV and ART (treatment as prevention, benefits of early ART); (ii) risk perception (morbidity and mortality); standard DHS measures on (iii) previous use of HIV services and (iv) self-rated health;^12^ and (v) WHO staging at enrollment.
- *Medical Chart Reviews:* Identifiers will be collected for all men enrolled in the study, including name, age, village and address, and phone number. Identifiers will be used to conduct medical chart reviews at 2-, 4-, 7-, and 12-months after enrollment as the primary measure of ART initiation and 6-month viral suppression (defined as an undetectable viral load of <200 copies/ml). Facility staff will review medical records at study facilities and all other Partners in Hope supported facilities within participating districts (61 facilities in total) to account for men who engage in ART outside study facilities. We successfully used this method in other HIVST studies to capture ART initiation.^7^ Male partners who are not found in medical chart reviews will receive a follow-up home-visit to confirm ART outcomes through review of their individual medical record book (health passport) and self-reporting in the event that there are gaps in the record. Men who cannot be reached will be counted as failures for true ART initiation. (Appendix I)
- *In-Depth Interviews:* In-depth interviews will be conducted with 120 randomly selected men and female partners to understand individuals’ experience with the interventions, contextual factors influencing ART engagement, and unintended consequences and adverse events related to the interventions.
- *Process Implementation Data:* The Implementation Team will keep daily logs as part of study monitoring and evaluation tools in order to assess the implementation of the intervention for each participant. Primary events to be recorded in the daily logs are: (1) unable to reach participant (and reason); (2) contacted participant; (3) intervention provided (and notes about the challenges and successes of the interaction; and (4) other comments relevant to intervention implementation. Each event will be recorded with a corresponding date.

Primary and secondary outcomes, along with their measurements, are described in Table 6.

***Table 6:*** *Study Measures for Aim 1*

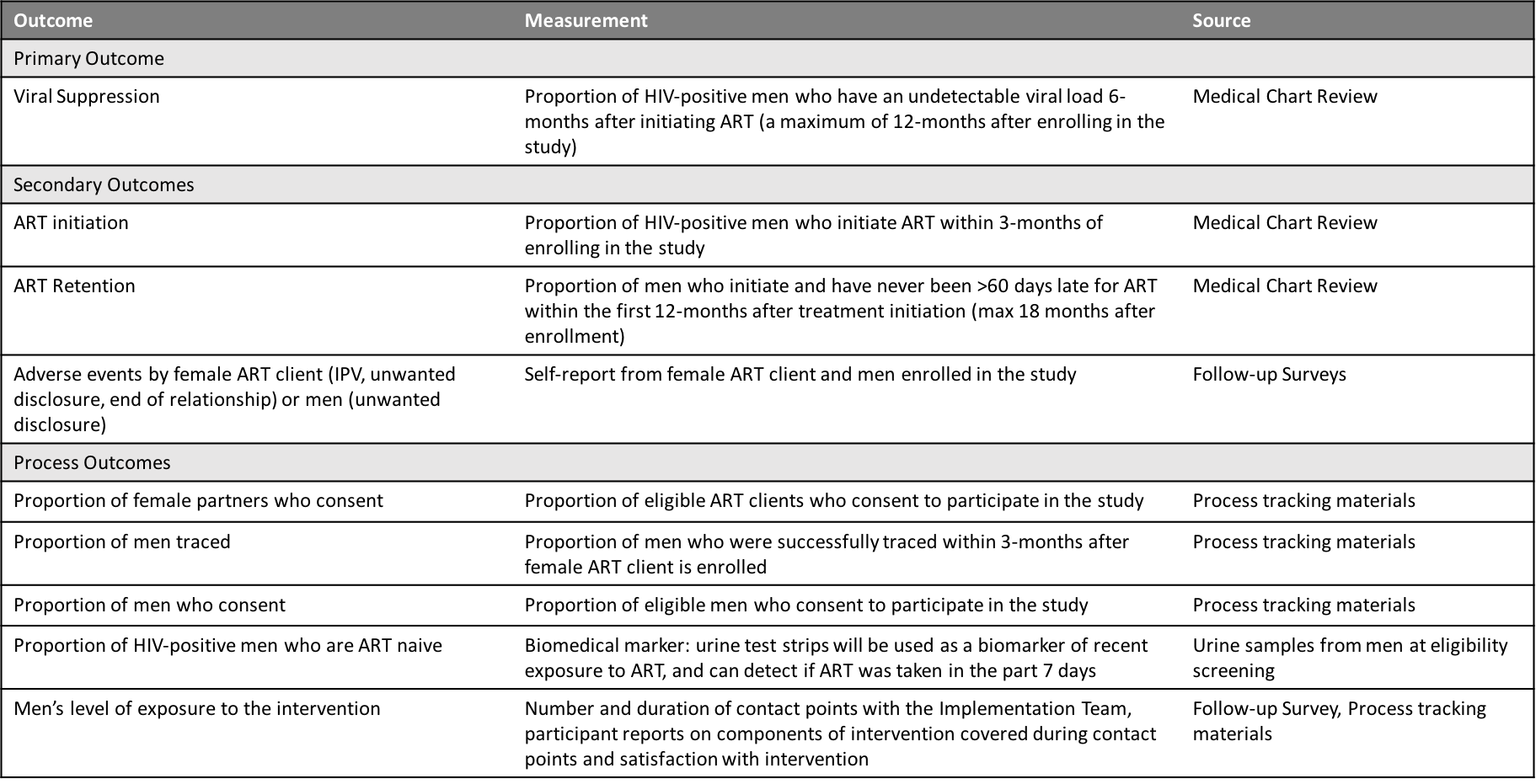

- *Sensitivity analyses for men excluded from the trial***:** We recognize that men’s consent to participate in the study may bias the sample enrolled in the study. There are two groups that we may not be able to include in the main study: (1) men we are unable to trace/contact (herein referred to as “unreachable men”) and (2) men we are able to reach but who refuse to participate in the main trial (herein referred to as “male refusers”). We will take two approaches to address this potential bias
  - Unreachable Men*:* We will collect data on men who are identified as potential respondents but unreachable via various tracing activities. Data will be collected from routine medical data (if identified through medical chart reviews) or from baseline interviews with their female partners (if identified through female partners). Data will include demographics, sexual partnerships and couple dynamics, and men’s history with health services, and HIV services specifically, as available.
  - Male Refusers*:* Men who are contacted but do not consent to the trial will be consented for a one-time survey immediately following refusal for the larger study. The same data will be collected, as described above.

##### 3.3.6. Sample size determination

***Aim 1***

We powered the study to detect differences in viral suppression between all arms and the Standard of Care arm at 12-months. Assuming that 50% of men in the Standard of Care arm and 70% of men in the intervention arms initiate ART and 60% of them reach viral suppression, and that 20% of men will be lost to follow-up in either arm (and treated as failures for outcome evaluation), the success proportions will be 0.240 and 0.336 in the various arms. The sample size needed to detect this difference with the power of 0.8 is 350 men per arm. The calculation is based on asymptotic normality of log odds ratio. We need to enroll and randomize a total of 700 HIV-positive men.

***Aim 2***

Survey sample size details provided in Aim 1. The number of interviews required for qualitative data can be challenging to predict. Data should be collected until saturation is reached, meaning that no new themes or relevant information is emerging. The exact number of interviews required to reach saturation differs based on the aim of the study, the diversity in respondents, and the theoretical framework used for analysis.^31^ However, a basic rule of thumb is that no sample size should be under 25 participants in order to reach saturation and identify all relevant themes or new information important to the study. We will enroll 30 individuals per in-depth interview category in order to ensure saturation.

##### 3.3.7. Data analysis

***Aim 1***

All randomized men will be included in the analysis of primary outcomes; men with missing outcome assessment due to loss to follow-up will be treated as outcome failures. All primary outcomes are binary; they will be analyzed by logistic regression models with intervention as a predictor, adjusted for baseline socioeconomic and demographic variables. We will conduct sensitivity analyses to account for men who we were never able to contact (unreachable men) and men who refused to participate in the full trial (refusers). We will run several analyses whereby the denominator includes (1) unreachable men and refusers; and (2) refusers.

***Aim 2***

*Surveys:* Subjects with complete data in outcomes as well as predictors will be included in the analysis. We will calculate descriptive statistics, including mean/median, variation (standard deviation, kurtosis), range, and frequency distributions for the demographic and clinical characteristics, overall and by study arm. Logistic models will be developed for the probability of a positive outcome, with sociodemographic factors included as covariates in a suitable form (linear/spline/factor). Differences in the prevalence of each of the outcomes of interest will be examined by study arm as well as by other factors of interest including demographic characteristics (e.g., age), couple characteristics, and knowledge/perceptions and biomedical knowledge. The differences will be evaluated using t-tests, Mann-Whitney U test (or other non-parametric tests), chi-square methods, and Fisher’s exact test as appropriate.

*In-depth interviews:* Audio recordings of in-depth interviews will be transcribed and translated to English. A preliminary codebook will be developed for both interview types (male and female). Selected investigators will a piloted codebook by independently reading and coding a randomly-selected subset of transcripts. Through an iterative consultative process, each investigator will revise their respective codebook and repeated this process until there was high interrater reliability among the group. All transcripts will be coded in Atlas.ti v8.3 using constant comparison, and coding disagreements were resolved by consensus.

#### 3.4. Aim 3

**Aim 3.** Determine the cost-effectiveness and scalability of hbART at a national level.

##### 3.4.1. Study design

We will conduct an incremental cost-effectiveness analysis and mathematical modelling to determine national scale-up potential. The average cost per successful outcome (early ART retention) will be calculated and compared across arms incrementally.

##### 3.4.2. Data collection techniques and tools

Costs will be measured from the health care provider. We will use micro-costing methods by first creating an inventory of all the resources used to achieve the observed study outcomes including:

- Male-focused counseling interactions (staff cadre, training received, duration of interaction and distance from facility travelled where applicable)
- Peer support interactions (staff cadre, training received, duration of interaction and distance from facility travelled where applicable)
- Provider interactions (staff cadre, training received, duration of interaction and distance from facility travelled where applicable)

For each study patient, the quantity (number of units) of resources used will be determined. Unit costs of resources, which are not human subject data, will be obtained from external suppliers and the site’s finance and procurement records and multiplied by the resource usage data to provide an average cost per study patient across centers in each study arm.

##### 3.4.3. Data analysis

**Cost-Effectiveness:** Using the average cost per patient as described above, we will then estimate the cost per outcome achieved in each arm. The main measure of effectiveness for the cost-effectiveness analysis will be both the primary study outcome (viral suppression at 6-months). We will calculate the difference in cost divided by the difference in effectiveness among study arms. Costs will be reported as means (standard deviations) and medians (IQRs) in USD, using the exchange rate prevailing during the follow up period.

**National scale-up modeling**: To determine the budget impact and affordability of the intervention arms, we will parameterize a national scale-up model using the study output. To determine the total cost and impact of the three intervention arms, as well as combinations of interventions, we will model cost and impact out to early ART retention (ART initiation and completion of the 4-week ART refill appointment). The following parameters to be estimated from this trial include:

- Proportion of men that initiate ART
- Proportion of men that complete the 4-week ART refill appointment
- Proportion of men that reached viral suppression at 6-months after ART initiation
- Proportion of men that are retained in care at 12-months after ART initiation

We will then estimate the expected increase in the number of men linked to ART after index HIVST, adjusted by facility type where possible, by each intervention arm. The number of facility-level HIV tests conducted through index testing at all 652 public healthcare facilities in Malawi from Oct 2019-Sept 2020 will be used for these national calculations. Each intervention will be tested separately in this model, as well as different combination of interventions. Different scenarios will be explored where interventions are used at different facilities (urban versus rural targeting of interventions, geospatial targeting of interventions), or different groups of men within the same facility (where data suggest that different demographics of men respond differently to the different interventions).

The national-level costs and expected number of men linked to ART, by each intervention and combinations of interventions, will be reported from this model. We will then contextualize the national cost of each intervention with a short-term 3-year budget impact: percent increase (or decrease) of the national HIV treatment budget with the inclusion of one of these interventions

### 4. ETHICAL CONSIDERATIONS

There is minimal risk associated with the above-mentioned procedures. We have extensive experience measuring ART initiation among men in Malawi. We conducted some of the first trials in the region to objectively measure ART initiation among men after receiving HIV self-testing through the Index HIVST Trial (PI: Dovel) and PASTAL Trial (male partners of antenatal clients; PI: Choko). We draw from lessons learned from our previous trials.

*Informed Consent*

Informed consent will be obtained before any study-specific procedures are performed. The informed consent process will include information exchange, detailed discussion, and assessment of understanding of all required elements of informed consent, including the potential risks, benefits, and alternatives to study participation. The process will emphasize the randomized nature of the study and the differences that participants may experience as part of the study relative to current local standards of care. The study will include children 15 years of age and older. Following Malawian protocol, adolescents <18 years of age will be required to attain assent before completing the survey. Based on prior studies, we anticipate <10% of participants to be under 18 years of age, providing a small sample size to explore the potential impact of facility-based testing for youth.

*Potential Benefits*

Men who participate in the study may have access to additional HIV services not usually provided through routine care, male specific counseling, motivational interviewing, and home-based ART initiation. Men can refuse HIV services at any point. Further, both men and women will have the opportunity to discuss their use of HIV services and any concerns with HIV. Information learned in this study may be of benefit to participants and others in the future, particularly information that may lead to optimized testing guidelines.

*Potential risks and discomforts*

Study procedures have minimal risk to the client. For men, maintaining privacy and confidentiality is a potential risk, particularly with home-based ART initiation. In our prior work delivering routine Index HIVST, we have had health workers visit a cluster of homes (not just one) to avoid unwanted questions about the individual’s serostatus. This has worked quite well, with no reports of unwanted disclosure, and we will use this approach in our proposed study to minimize risk of unwanted disclosure. Further, men may refuse any ART service at any point if they are uncomfortable.

For women, increased intimate partner violence (IPV) may be a potential risk, particularly if their male partner is prone to violence. To reduce these risks, women who report IPV with their current partner in the past 12 months will be excluded from the study. Female ART clients who report IPV at any point of the intervention will be withdrawn from the study, along with their male partner, counseled, and referred to community-based resources for IPV. We also will provide extensive counseling on status disclosure and an IPV hotline to all female participants. Further, Our PASTAL and Index HIVST Trials show no sign of increased IPV and we have published extensively on risk factors for IPV in other settings.^71-73^

Finally, Participation includes completion of a survey that will assess previous use of health services, perceptions of health services received, and sociodemographic and biomedical factors that may be associated with health service utilization. Participants may feel some psychological stress or discomfort from some of the questions, although most questions are not sensitive in nature. Participants may decline to answer any questions that make them uncomfortable and may end participation at any time.

Reimbursement/compensation

Participants will be provided MK 7,500 (equivalent to 10USD) for each survey completed (male participants: MK 30,000 / 40USD across the duration of the study; female participants: MK 30,000 / 40USD across the duration of the study). They will receive the above compensation regardless if they use HIV services or not. Those who complete the additional in-depth interview 6-months after study enrollment will receive an additional MK 7,500 (equivalent to 10USD) for their time.

Privacy and confidentiality

All study procedures will be conducted in private, and every effort will be made to protect participant privacy and confidentiality to the extent possible. Participant information will not be released without written permission to do so except as necessary for review, monitoring, and/or auditing. All study-related information will be stored securely. Participant research records will be stored in locked areas with access limited to study staff. All study data will be identified by participant ID (PID) only. Likewise, communications between study staff and protocol team members regarding individual participants will identify participants by PID only. Process evaluation documents, such as intervention monitoring and evaluation tools, will only include PID and will not store PID and identifiers together. All local databases will be encrypted and secured with password-protected access systems. Lists, logbooks, appointment books, and any other documents that link PID numbers to personal identifying information will be stored in a separate, locked location in an area with limited access. For the intervention, home visits will be conducted by health workers who visit a cluster of homes (not just one) at one time in order to avoid unwanted questions about the individual’s serostatus. This has been used in other interventions focused on partner testing and treatment with high success of removing unwanted disclosure to community members.

### 5. DISSMINATION OF RESULTS

This study will set the stage for interventions that combine HIVST with differentiated models for early ART retention in low-resource settings. The study is timely and of high-impact. Findings will establish the effectiveness of home-based ART among men, and can directly inform HIV programs throughout the region. The dissemination plan was developed to achieve the most impact while still ensuring dissemination among local stakeholders who may immediately benefit from study findings.

Partners in Hope is already integrated into national technical working groups, so dissemination will follow standard meeting schedules and draw upon Partners in Hope’s longstanding history with the Ministry of Health. Additionally, we will disseminate results through presentations at international scientific meetings and through high-impact peer-reviewed journals. The mentorship team has extensive experience publishing in high-impact journals (e.g., *AJPH, AIDS, BMJ, Lancet HIV, JAIDS*, *PLOS Med*)

### 6. PERSONNEL ROLES AND INSTITUTIONS

The proposed research team includes clinical researchers and implementation science professionals with substantial experience in HIV testing, HIV prevention and treatment, cost effectiveness, differentiated care model studies, and male-focused studies and programs in Malawi and Sub-Saharan Africa. The study will be implemented in partnership with Partners in Hope Medical Center in Lilongwe, which has years of experience collaborating with Ministry of Health and local health facilities on similar studies, mentoring staff, and running studies embedded within routine clinical care.

- Thomas Coates, PhD, Principle Investigator, Division of Infectious Disease UCLA
- Augustine Choko, PhD, Site Principle Investigator, Malawi Liverpool Wellcome Trust
- Kathryn Dovel, MPH, PhD, Principle Investigator, Division of Infectious Disease University of California Los Angeles (UCLA) and Research Director for Partners in Hope
- Risa Hoffman, MPH, MD, Co-Investigator, Division of Infectious Disease UCLA
- Michal Kulich, Biostatistician, Charles Univeristy in Prauge
- Misheck Mphande, MPH, Study Coordinator, Partners in Hope
- Julie Hubbard, MSc, Study Coordinator, Division of Infectious Disease UCLA
- Kelvin Balakasi, Study Data Manager, Parners in Hope
- Khumbo Phiri, Implementation Science Manager, Partners in Hope

CV’s for participating personnel are provided in the Appendix L.

### 7. REGULATORY OVERSIGHT

This study is sponsored by the National Institute of Mental Health and implemented through Partners in Hope (PIH), Malawi. PIH staff will perform monitoring visits. As part of these visits, monitors will inspect study-related documentation to ensure compliance with all applicable regulatory requirements.

All health facilities will receive an Initial Registration Notification from PIH that indicates successful completion of the protocol registration process. A copy of the Initial Registration Notification will be retained in the site's regulatory files.

We have developed a trial advisory group. See Table 6 for details about the group members. The group will meet every quarter to review progress, and challenges with study implementation, and provide input on findings.

***Table 6.*** *Description of trial advisory group*

| **Name** | **Affiliation** | **Expertise** |
| --- | --- | --- |
| Dr. Morna Cornell | University of Cape Town | Epidemiologist, health system barriers to men’s care, men’s HIV services, advocacy and policy change |
| Dr. Heidi van Rooyen | SA Human Sciences Research Council | Social scientist, HIV vulnerability and inequality, interventions for men’s ART initiation |
| Dr. Deborah Donnell | University of Washington, Fred Hutch Vaccine and Infectious Disease Division | Biostatistician, international HIV trials, PI of the HPTN Statistical and Data Management Center |
| Dr. Connie Celum | University of Washington | Infectious disease physician and epidemiologist, implementation science in Africa, HIV prevention trials |
| Dr. Thoko Kalua | Malawi Ministry of Health, Deputy Director at Department of HIV and AIDS | Epidemiologist. Extensive experience in national HIV programs, M&E, and scale-up of interventions on the ground |
| Dr. Sergio Chicumbe | Mozambique National Health Institute (INS), Health System Research Cluster | Clinical trials and implementation science. Extensive experience in national public health programs, methodology for health services research and quality care improvement. |

For any future protocol amendments, upon receiving final IRB/EC and any other applicable regulatory entity approvals, sites should implement the amendment immediately. Sites are required to submit an amendment registration packet to the PIH Protocol Team. PIH key personnel will review the submitted protocol registration packet to ensure that all the required documents have been received.

### 8. STUDY IMPLEMENTATION

Study implementation at each site will be guided site-specific standard operating procedures (SOPs). These SOPs will be updated and/or supplemented as needed to describe roles, responsibilities, and procedures for this study.

### 9.  PROTOCOL DEVIATION REPORTING

All protocol deviations will be documented in participant research records. Reasons for the deviations and corrective and preventive actions taken in response to the deviations will also be documented. Deviations will be reported to site IRBs/ECs and other applicable review bodies in accordance with the policies and procedures of these review bodies. Serious deviations that are associated with increased risk to one or more study participants and/or significant impacts on the integrity of study data must also be reported to the Protocol Team as soon as possible.

### 10.  WORK PLAN TIMELINE

***Table 7:*** *Anticipated work plan timeline of study activities, by year*
