## Supporting Information S3 for "Engaging Men Through HIV Self-Testing with Differentiated Care to Improve ART Initiation and Viral Suppression among Men in Malawi (ENGAGE): a study protocol for a randomized control trial"

UCLA OHRPP
Office of the Human Research Protection Program

University of California Los Angeles
10889 Wilshire Blvd, Suite 830
Los Angeles, CA 90095-1406

http://ora.research.ucla.edu/ohrpp
General Campus IRB: (310) 825-7122
Medical IRB: (310) 825-5344

### EXTERNAL REVIEW ACCEPTED

| | |
|---|---|
| **DATE:** | 6/25/2021 |
| **TO:** | THOMAS COATES, PhD<br>MEDICINE-INFECTIOUS DISEASE |
| **FROM:** | Principal Analyst, UCLA OHRPP |
| **RE:** | IRB#21-000594<br>Engaging Men Through HIV Self-Testing with Differentiated Care to Improve ART Initiation and Viral Suppression among Men in Malawi (ENGAGE) |

This letter documents UCLA's reliance on an external IRB's review for the above referenced study. UCLA's Federalwide Assurance (FWA) with the Department of Health and Human Services is FWA00004642.

**Registration Details:**

| | |
|---|---|
| Reviewing IRB | National Health Sciences Research Committee (NHSRC) |
| Type of Reliance | IAA |
| Protocol ID of the Reviewing IRB | Protocol # 20/07/2562 |
| Approval period of the Reviewing IRB | 7/17/2020-7/16/2021 |
| Acceptance date by UCLA | 6/25/2021 |
| Funding Source(s) | 1) NIH - NATIONAL INSTITUTES OF HEALTH<br>*Grant PI*: THOMAS COATES<br>*Grant Title*: Combining HIV self-testing with differentiated care models to increase ART initiation and viral suppression among men in Malawi<br>*Grant Number*: PA-19- 042 |
